# Differential Impacts of Perceived Social Support on Alcohol and Cannabis Use in Young Adults: Lessons from the COVID-19 Pandemic

**DOI:** 10.1101/2022.08.04.22278446

**Authors:** Michelle J. Blumberg, Lindsay A. Lo, Geoffrey W. Harrison, Alison Dodwell, Samantha H. Irwin, Mary C. Olmstead

## Abstract

Coronavirus (COVID-19) lockdowns provided a unique opportunity to examine how changes in the social environment impact mental health and wellbeing. We addressed this issue by assessing how perceived social support across COVID-19 restrictions alters alcohol and cannabis use in emerging adults, a population vulnerable to adverse outcomes of substance use. Four hundred sixty-three young adults in Canada and the United States completed online questionnaires for three retrospective time points: Pre-Covid, Lockdown and Eased Restrictions. Sociodemographic factors, perceived social support, and substance use were assessed. Overall, alcohol use decreased while cannabis use increased during Lockdown. Interestingly, social support negatively predicted alcohol use and positively predicted cannabis use during Lockdown. These findings suggest a difference in motives underlying alcohol and cannabis use in emerging adults. Importantly, these changes were not sustained when restrictions eased, suggesting that emerging adults exhibit resiliency to the impacts of COVID-19 restrictions on substance use.

## Introduction

The coronavirus (COVID-19) was declared a pandemic by the World Health Organization in March 2020 (World Health Organization, 2020). Following this, efforts to limit spread of the virus included widespread stay at home orders, and the closure of non-essential services, schools, and workplaces. This led to immense changes in daily life that were particularly impactful on young adults who rely on social interactions and social support for wellbeing (Asher & Weeks, 2013; Sandstrom & Dunn, 2014; Dissing et al., 2019; Richards et al., 2019; Scardera et al., 2020). Social support is broadly defined as the assistance and protection given to others (Shumaker & Brownell 1984, Wortman & Dunkel-Schetterm, 1987), and consists of four defining attributes: emotional, instrumental, informational, and appraisal support (Barrera, 1986). Constituents to social support include one’s social network, which is the interactive field of persons that provide helpfulness and protection (Berkman, 1984, Gottlieb, 1983), and social embeddedness, which is the connectedness people have to others within their social network (Barrera 1986, Langford et al 1997). Concerns over the impact of COVID-19 on mental health (Abbott, 2021; Galea et al., 2020; Singh et al., 2020), specifically substance use (Czeisler et al., 2020; Ornell et al., 2020), have simultaneously increased. Consequently, government mandated lockdowns provided a unique opportunity to assess the impact of social support on substance use. The goal of the current study was to integrate these two areas by investigating the impact of social support on alcohol and cannabis use among young adults in Canada and the United States throughout the COVID-19 pandemic.

Alcohol and cannabis are two of the most widely used psychoactive substances (Peacock et al., 2017; Vincenzi et al., 2017), and are both associated with acute and chronic negative effects. Generally, studies have found increased alcohol and cannabis use during COVID-19 in adult samples (e.g., Taylor et al., 2021; Barbosa et al., Dumas et al., 2020; 2020; Jacob et al., 2020; Grossman et al., 2020; Sharma et al., 2020; Wardell et al., 2020), but these patterns may differ in young adult samples. Young adulthood is generally accepted in the psychological literature to refer to the crucial developmental period following adolescence (emphasizing identity formation) that most individuals reach between the ages of 19 and 39 years (Erikson, 1965). Critically, the literature investigating alcohol and cannabis use patterns in young adults during the COVID-19 pandemic is much slimmer, and these studies have obtained mixed results. For example, both increases (Romm et al., 2021; Sharma et al., 2020; Lechner et al., 2020) and decreases in alcohol consumption have been reported in young adults (Bollen et al., 2021; White et al., 2020). Of the available literature on changes in cannabis use, no change and increases in use have both been reported (Romm et al., 2021; Graupensperger et al., 2021; Bonar et al., 2021; Papp & Kouros, 2021). Furthermore, there is a lack of research directly comparing patterns of alcohol and cannabis use in this population across the pandemic. These contradictory findings may reflect differences in how substance use was assessed and alterations in social restrictions associated with different phases of the pandemic. Identifying the factors that impact patterns of alcohol and cannabis use in young adults during COVID-19 is important as this group is particularly vulnerable to developing substance use disorder and other mental health conditions, including both internalizing (characterized by anxiety, depression, and somatic symptoms) and externalizing (characterized by impulsive and disruptive symptoms) disorders (American Psychiatric Association, 2013; Brewer et al., 2017; Gray & Squeglia, 2017; Hamidullah et al., 2020).

Contradictory findings regarding substance use during COVID-19 suggest that individual factors impacted alcohol and cannabis use in young adults during this period. Given the dramatic changes in social interactions during lockdowns, differences in social support may explain the conflicting evidence obtained from previous studies. Evidence supports that social support encourages positive health behaviors and is also a stress-buffering mechanism (Emmons et al., 2007, Wills et al., 2004, Wills & Ainette, 2012). As such, social support is well supported in the literature to be associated with substance use (Wills et al., 2004, Wills & Ainette, 2012). More specifically, social integration and social support are linked to decreased prevalence of smoking and heavy drinking (Brennan & Moss, 1990; Romano, Bloom, & Syme, 1991). Social support may also mitigate the risk of problem drinking in young adults through reduced psychological distress (Segrin et al., 2016). Lower familial support is associated with increased substance use among young people (Piko et al., 2000; Beitchman et al., 2005; Arias-De la Torre, 2019). Therefore, individual differences in social support may explain divergent findings in substance use patterns during COVID-19.

Lower social support and related factors have been associated with increased substance use during COVID-19 in adults (Wardell et al., 2020; Vanderbruggen et al., 2020; Clair et al., 2020; Horigian et al., 2020), but findings in younger adult samples are less clear. Some studies have replicated a negative relationship between social support-related factors and drinking (Lechner et al., 2020; Sharma et al., 2020; Bollen et al., 2021). Although few studies have examined the relationship between social support and cannabis use in young adults during COVID-19, two studies suggested factors related to social support (e.g., loneliness, isolation) may also be associated with increased cannabis use (Bonar et al., 2021; Bartel et al., 2020). There is a growing interest in investigating the impact of social support (particularly loneliness and living alone) on substance use among young adults (e.g., Horigian et al., 2021; Sylvestre, 2022). However, further research is needed to understand the direct impact of perceived social support from family, friends, and significant others on alcohol and cannabis use throughout different periods of COVID-19 restrictions.

Accordingly, the goal of the current study was to investigate changes in alcohol and cannabis use in young adults across three phases of the pandemic. Timepoint 1 refers to the period preceding the pandemic onset. Timepoint 2 refers to the first wave of COVID-19 (i.e., tight restrictions that limited in-person interactions). Timepoint 3 refers to the period after the first wave (i.e., ease of restrictions). Additionally, we sought to investigate the ways in which perceived social support impacted substance use among young adults throughout these phases. First, we hypothesized that alcohol and cannabis use would increase during the first wave of COVID-19 when lockdown orders were in place. Second, we hypothesized that perceived social support would modulate changes in substance use throughout COVID-19. Specifically, we predicted that only those with low levels of social support would experience an increase in substance use during the first wave.

Concerns of the impact of the COVID-19 pandemic on health outcomes, specifically mental health, have persisted. Given that young adulthood is a vulnerable period for developing mental health conditions, including substance dependence, findings from this study provide valuable information on how social restrictions impact substance misuse and may help guide preventative or intervention measures in the future.

## Methods

### Sample and Design

This study used a longitudinal retrospective survey design. Five hundred fifty-two adults (19-30 years of age) were recruited from Canada and the United States via social media (e.g., Facebook, emails listservs, Instagram, Twitter) and the Queen’s University participant pool. Authors (MJB, LAL, AD, and SHI) distributed the survey link and study advertisement via these platforms. To screen for eligibility, participants were first asked to indicate their country of residence, their age, and whether they had consumed alcohol and/or cannabis in 2020. Those who were not living in Canada or the United States, were not between 19 and 30 years of age, and who did not consume alcohol and/or cannabis at any point in 2020 were excluded from the study. The same participants completed a survey (approximately 15 minutes) that gathered information on sociodemographic characteristics, perceived social support, and substance use at three distinct retrospective time points. Data were collected between August and November 2020 using the following time points: Timepoint 1 = January to early March 2020 (Pre-COVID); Timepoint 2 = mid-March to the end of May 2020 (Lockdown); and Timepoint 3 = June to July 2020 (Eased Restrictions). The questionnaire was designed and administered through Qualtrics Survey Software (Qualtrics, Provo, UT). Respondents were either entered in a draw to win one of ten Amazon or Starbucks gift cards valued at $25 each or received course credit for their participation. Incomplete surveys were excluded. For this analysis, the small number of individuals reporting unemployment before COVID-19 (n = 11) were excluded due to higher reported levels of substance use compared to students and employed young adults, as meaningful conclusions cannot be made due to the small group size. The final sample was 463.

### Measures

Sociodemographic variables included gender, age, completed level of education, and employment status at the time of survey completion.

#### Perceived social support

Social Support was measured at each time point using the Multidimensional Scale of Perceived Social Support (MSPSS, Zimet et al., 1988). The MSPSS is a validated scale that measures subjective feelings of social support from family, friends, and significant others. The MSPSS includes 13 questions rated on a 7-point Likert scale (1 = Very strongly disagree to 7 = Very strongly agree). Some example items are “*I could talk about my problems with my family” and* “*I could count on my friends when things went wrong”*. Items were summed to obtain a composite Social Support score for each respondent.

#### Alcohol and Cannabis Use

Alcohol and cannabis use were measured using a variation of the Timeline Followback Alcohol and Marijuana Use Calendar, a well-validated calendar assisted alcohol and cannabis use measure, for up to 12 months prior (TLFB, Sobell & Sobell, 1992; Sobell et al., 2001). Specifically, respondents were asked to provide retrospective estimates of the number of days alcohol was consumed and the number of alcoholic drinks consumed in a typical week for each time point. Respondents were then asked to report the number of days and the number of times cannabis was ingested during a typical week within each time point. More specifically, respondents were told to consider one ingestion of cannabis to be one occasion in which they consumed Tetrahydrocannabinol (THC)-containing cannabis products, such as one joint, pipe or an edible. Number of days was used as our measure of frequency of alcohol and cannabis consumption. Number of drinks (alcohol) and ingestions (cannabis) was used as indicators of the quantity of alcohol and cannabis consumed.

#### Statistical Analysis

All analyses were conducted on the four dependent measures of substance use: frequency of alcohol consumption (number of days/week), quantity of alcohol consumed (number of alcoholic drinks/week), frequency of cannabis consumption (number of days/week), and quantity of cannabis consumed (number of ingestions/week). Initial one-way repeated measures Analysis of Variances (ANOVAs) were run to determine the impact of time points on Social Support and levels of substance use. Following this, four linear mixed-effects models were used to predict each substance use measure from Social Support at each of three COVID time points coded as fixed effects, and participants coded as random effects (Kristensen and Hansen, 2004; Magezi, 2015). Linear mixed-effects models are a more appropriate analytic tool for repeated measure designs than repeated measures ANOVAs as they accommodate missing data, are more likely to meet their underlying assumptions, provide better interpretability, model variance at the participant level, and provide more accurate parameter estimates (Kristensen and Hansen, 2004). The three time points of COVID were dummy coded with Lockdown as the reference, creating two dummy coded variables. Dummy coded variable one represented a comparison between Pre-COVID and Lockdown and dummy coded variable two represented a comparison between Eased Restrictions and Lockdown. Social Support was grand mean centered. The full model included both dummy coded COVID time points, grand mean centered Social Support, and their interactions with gender and level of education included as covariates. For the purposes of following up significant interactions, we examined Social Support at mean levels (moderate Social Support) and at 1 SD above (higher Social Support) and 1 SD below (lower Social Support) mean levels. Statistical analyses were the lmer package (Bates et al., 2014) carried out using R (version 3.6.2) and RStudio (version 1.2.5033).

## Results

### Sample Characteristics

Table 1 presents sample characteristics of participations. Scores on the MSPSS ranged from 12 to 84 (i.e., low to high Social Support).

**Table 1.**
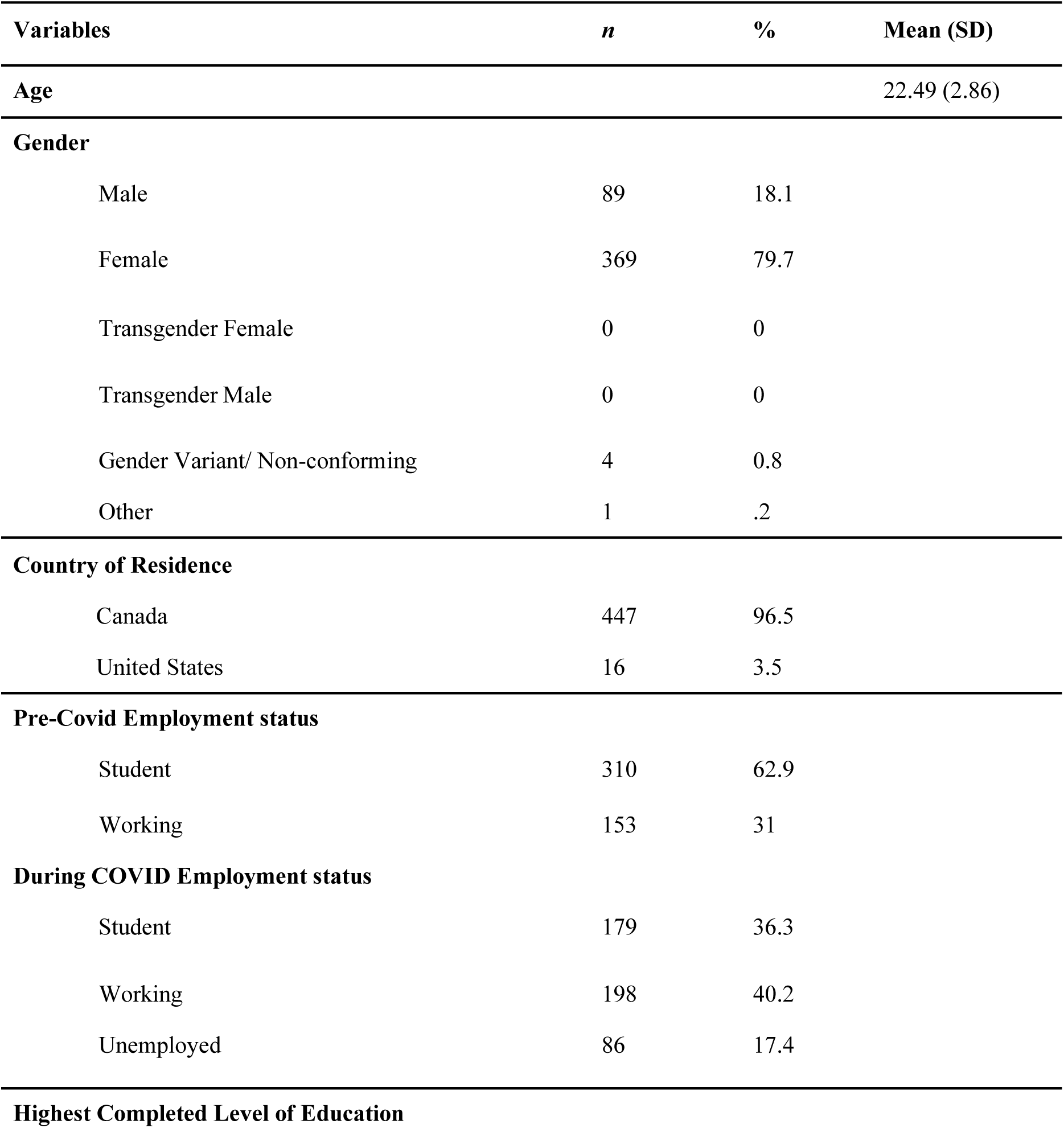
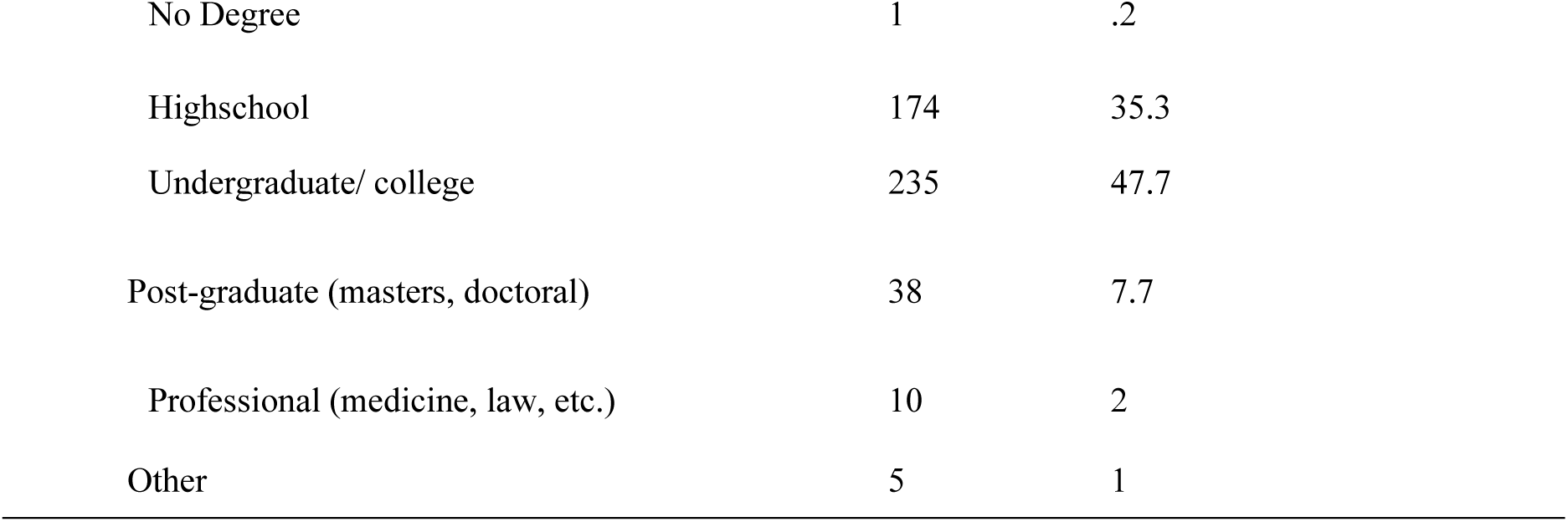
Sample Characteristics

| Variables | *n* | % | Mean (SD) |
| --- | --- | --- | --- |
| Age |  |  | 22.49 (2.86) |
| **Gender** |  |  |  |
| Male | 89 | 18.1 |  |
| Female | 369 | 79.7 |  |
| Transgender Female | 0 | 0 |  |
| Transgender Male | 0 | 0 |  |
| Gender Variant/ Non-conforming | 4 | 0.8 |  |
| Other | 1 | .2 |  |
| **Country of Residence** |  |  |  |
| Canada | 447 | 96.5 |  |
| United States | 16 | 3.5 |  |
| **Pre-Covid Employment status** |  |  |  |
| Student | 310 | 62.9 |  |
| Working | 153 | 31 |  |
| **During COVID Employment status** |  |  |  |
| Student | 179 | 36.3 |  |
| Working | 198 | 40.2 |  |
| Unemployed | 86 | 17.4 |  |
| **Highest Completed Level of Education** |  |  |  |

|  |  |  |
| --- | --- | --- |
| No Degree | 1 | .2 |
| Highschool | 174 | 35.3 |
| Undergraduate/ college | 235 | 47.7 |
| Post-graduate (masters, doctoral) | 38 | 7.7 |
| Professional (medicine, law, etc.) | 10 | 2 |
| Other | 5 | 1 |

### Overall Trends in Substance Use

Individuals’ level of Social Support decreased from Pre-COVID (*M* = 67.31, *SD* = 13.37) to Lockdown (*M* = 64.17, *SD* = 15.24), then increased again during Eased Restrictions (*M* = 65.94, *SD* = 15.36), *F*(2, 924) = 25.70, *p* < .001, *η_p_*^2^ = .053. The number of days Alcohol was consumed decreased from Pre-COVID (*M* = 2.01, *SD* = 1.65) to Lockdown (*M* = 1.34, *SD* = 2.49), then increased again during Eased Restrictions (*M* = 2.33, *SD* = 2.36), *F*(2, 924) = 38.31, *p* < .001, *η_p_*^2^= .077 (see Figure 2a). The quantity of Alcohol consumed decreased from Pre-COVID (*M* = 7.25, *SD* = 8.01) to Lockdown (*M* = 3.37, *SD* = 9.29), then increased again during Eased Restrictions (*M* = 5.75, *SD* = 8.17), *F*(2, 924) = 39.43, *p* < .001, *η_p_*^2^= .079.

The number of days Cannabis was consumed increased from Pre-COVID (*M* = 1.64, *SD* = 2.75), to Lockdown (*M* = 2.36, *SD* = 2.10), then decreased during Eased Restrictions (*M* = 1.66, *SD* = 2.75), *F*(2, 924) = 23.02, *p* < .001, *η_p_*^2^= .047. The quantity of Cannabis consumed increased from Pre-COVID (*M* = 3.96, *SD* = 10.11), to Lockdown (*M* = 6.56, *SD* = 8.80), then decreased during Eased Restrictions (*M* = 4.19, *SD* = 10.39), *F*(2, 924) = 19.35, *p* < .001, *η_p_*^2^= .040.

### Relationship between Substance Use and Social Support

#### Alcohol

##### Frequency of Alcohol Consumption

In order to determine the impact of Social Support during three COVID time points on alcohol consumption behavior, we ran a linear mixed-effects model predicting frequency of Alcohol consumption from the interaction between time point and Social Support. The model revealed significant interactions between both dummy coded COVID time point variables and Social Support, *b*s > 0.03, *t*s > 3.20, *p*s ≤ .001 (see Figure 1).

**Figure 1.**
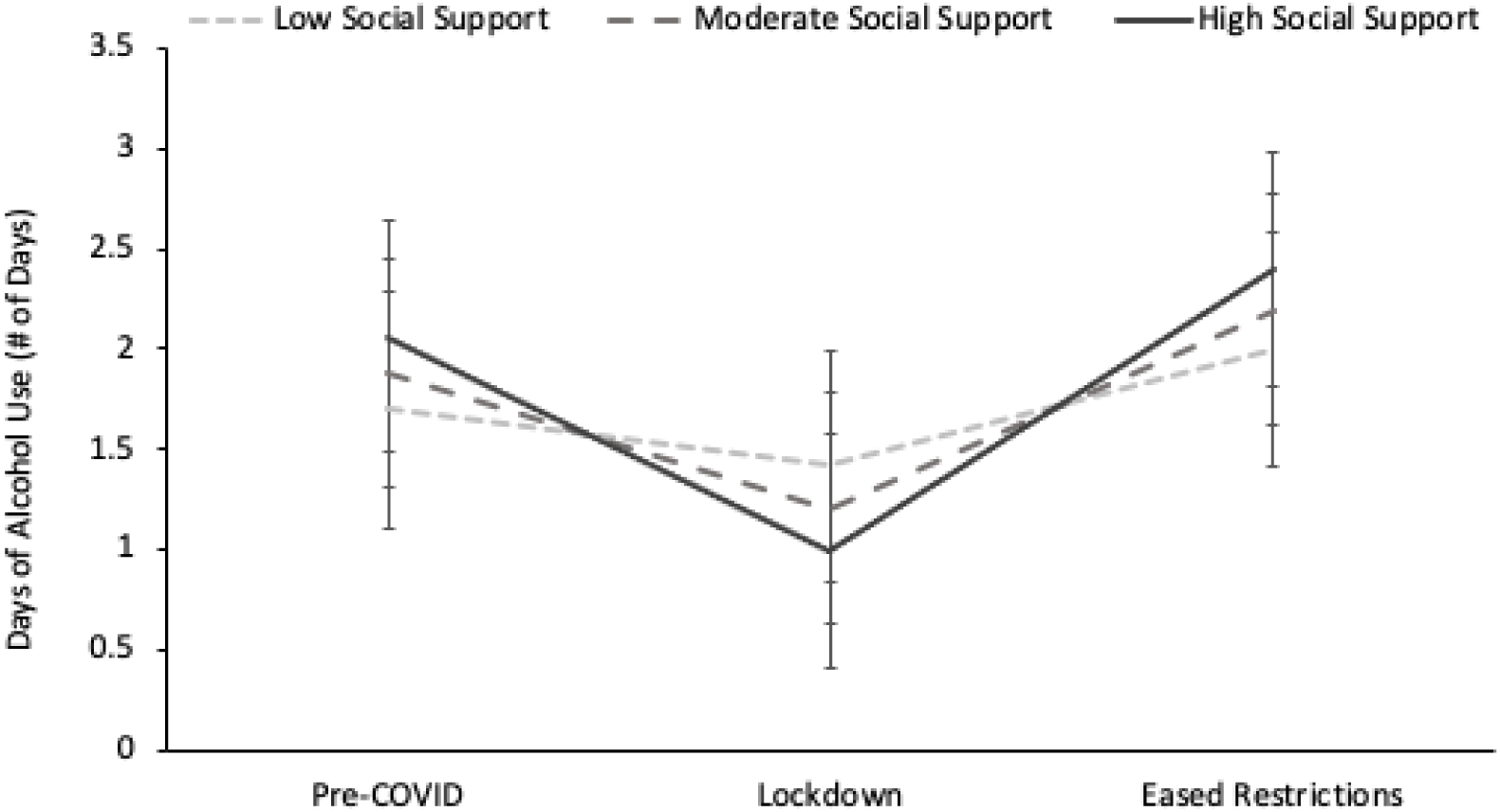
Changing relationship between social support and frequency of alcohol consumption throughout COVID-19 time points. Error bars show standard error of the means.

To follow up these interactions we first examined the relationship between Social Support and frequency of Alcohol use at each time point. Pre-COVID, frequency of Alcohol use was not related to Social Support, *b*(1357) = 0.013, CI = [-0.002, 0.027]. During Lockdown, Social Support negatively predicted frequency of Alcohol use, *b*(1347) = -0.014, 95% CI = [-0.027, -0.002]. During Eased Restrictions, Social Support positively predicted frequency of Alcohol use, *b*(1346) = 0.014, 95% CI = [0.001, 0.026].

Second, we examined differences in the frequency of Alcohol consumption across time points at each level of Social Support. For individuals with low Social Support, the frequency of Alcohol consumption was consistent between Pre-COVID and both Lockdown and Eased Restrictions, but significantly increased from Lockdown to Eased Restrictions, *t*(928) = -3.67, *p* = 0.001. For individuals with moderate to high levels of Social Support there was a significant decrease in the frequency of Alcohol use from Pre-COVID to Lockdown, all *t*s > 5.80, all *p*s < .001, and a significant increase in the frequency of Alcohol use from Lockdown to Eased Restrictions, all *t*s > 8.49, all *p*s < .001. However, only those with moderate Social Support reported significantly higher frequency of Alcohol use during Eased Restriction compared to Pre-COVID, *t*(916) = 2.73, all *p* = .018.

##### Quantity of Alcohol Consumption

Similar to the frequency of Alcohol consumption, when predicting the quantity of Alcohol consumption, both time point variables significantly interacted with Social Support, *b*s > 0.067, *t*s > 2.19, *p*s < .029 (see Figure 2).

**Figure 2.**
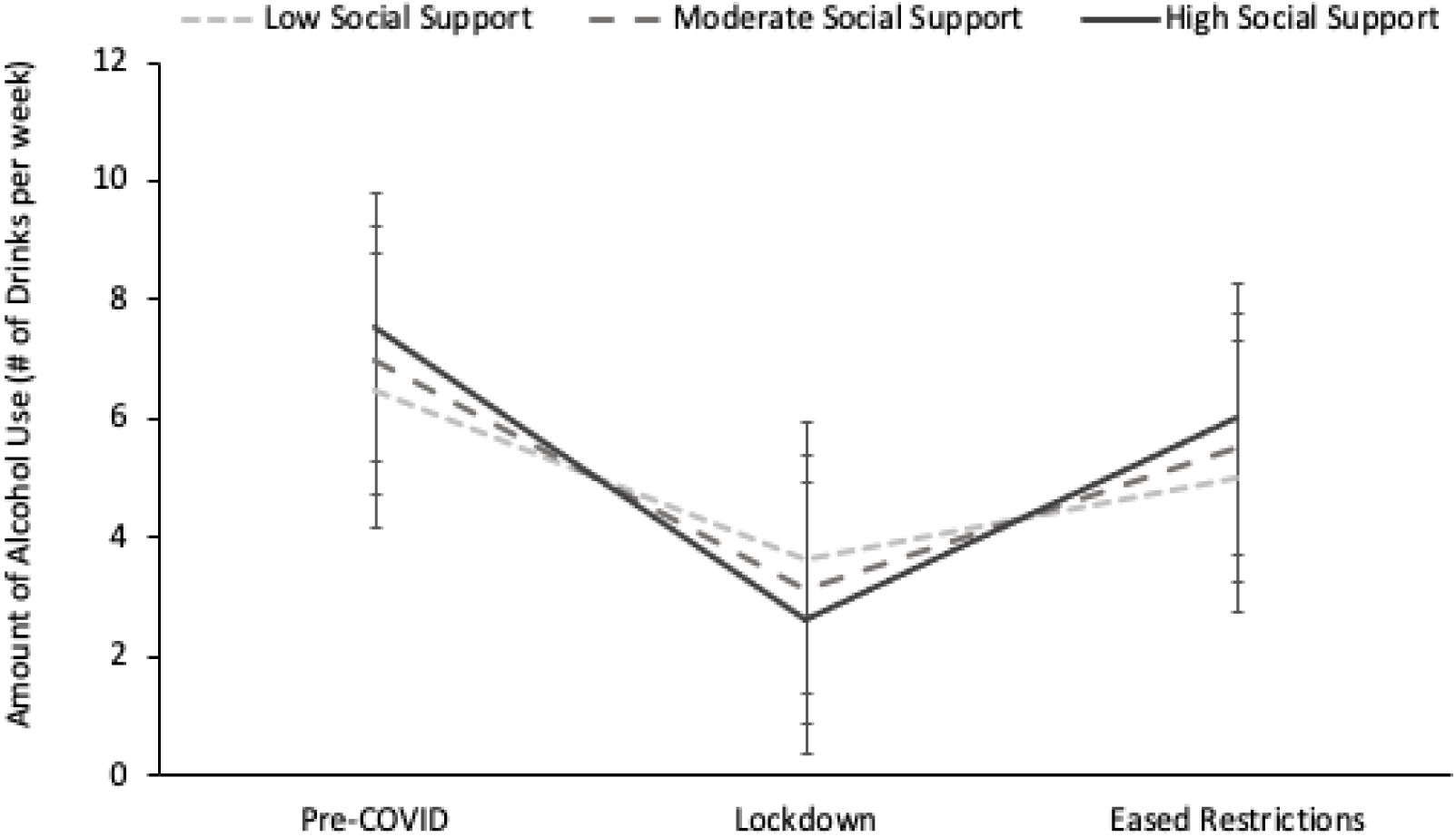
Changing relationship between social support and quantity of alcohol consumption throughout COVID-19 time points. Error bars show standard error of the means.

To follow up these interactions we first examined the relationship between Social Support and quantity of Alcohol use at each time point. Surprisingly, unlike for the frequency of Alcohol use, Social Support was not significantly related to the quantity of Alcohol consumed at any time point.

Second, we examined differences in the frequency of Alcohol consumption across time points at each level of Social Support. For all levels of Social Support, the quantity of Alcohol use decreased from Pre-COVID to Lockdown, all *t*s > 4.31, all *p*s < .001. For those with moderate and high Social Support, the quantity of Alcohol use increased from Lockdown to Eased Restrictions, all *t*s > 5.28, all *p*s < .001, whereas those with low Social Support saw a marginal increase, *t*(928) = 2.29, *p* = .06. Compared to Pre-COVID levels, individuals across all levels of Social Support saw at least a marginal drop in the quantity of Alcohol use during Eased Restrictions, all *t*s > 2.16, all *p*s < .079.

#### Cannabis

##### Frequency of Cannabis Consumption

In order to determine the impact of Social Support during three COVID time points on cannabis consumption behavior, we ran a linear mixed-effects model predicting the frequency of Cannabis consumption from the interaction between COVID time points and Social Support. Like with the frequency of Alcohol consumption, the model revealed significant interactions between both dummy coded time point variables and Social Support, *b*s > -0.04, *t*s > -5.10, *p*s < .001 (see Figure 3). However, the nature of the relationship between Social Support and Cannabis use during each COVID time point was very different.

**Figure 3.**
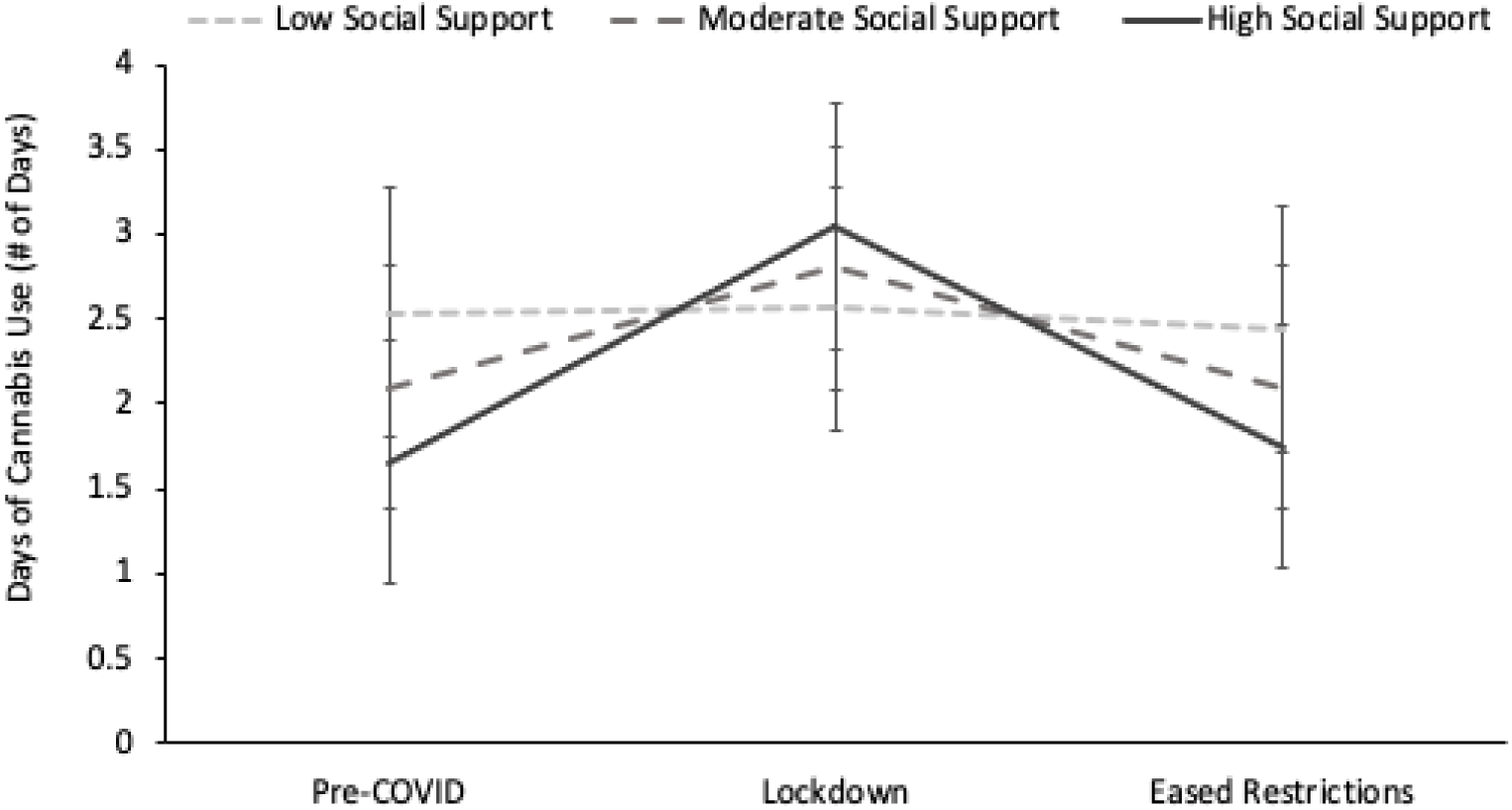
Changing relationship between social support and frequency of cannabis consumption throughout COVID-19 time points. Error bars show standard error of the means.

To follow up these interactions we first examined the relationship between Social Support and frequency of Cannabis use at each time point. Pre-COVID and during Eased Restrictions, Social Support negatively predicted the frequency of Cannabis consumption, all *b*s > -0.023, CI = [-0.037, -0.010]. In contrast, during Lockdown Social Support had the opposite effect, positively predicting the frequency of Cannabis use, *b*(1353) = 0.017, 95% CI = [0.003, 0.031].

Second, we examined differences in the frequency of Cannabis consumption across time points at each level of Social Support. For individuals with low Social Support, Cannabis use remained constant across all three COVID time points, all *t*s < 0.72, all *p*s > .75. For those with moderate and high Social Support, frequency of use increased from Pre-COVID to Lockdown, all *t*s > 5.85, all *p*s < .001, and then decreased from Lockdown to Eased Restrictions, all *t*s > 5.94, all *p*s < .001. The decrease during Eased Restrictions was a return to Pre-COVID levels, all *t*s < 0.53, all *p*s > .86.

##### Quantity of Cannabis Consumption

Similar to the frequency of Cannabis consumption, when predicting the quantity of Cannabis consumption, both time point variables interacted with Social Support, *b*s > -0.116, *t*s > 3.80, *p*s < .001 (see Figure 4).

**Figure 4.**
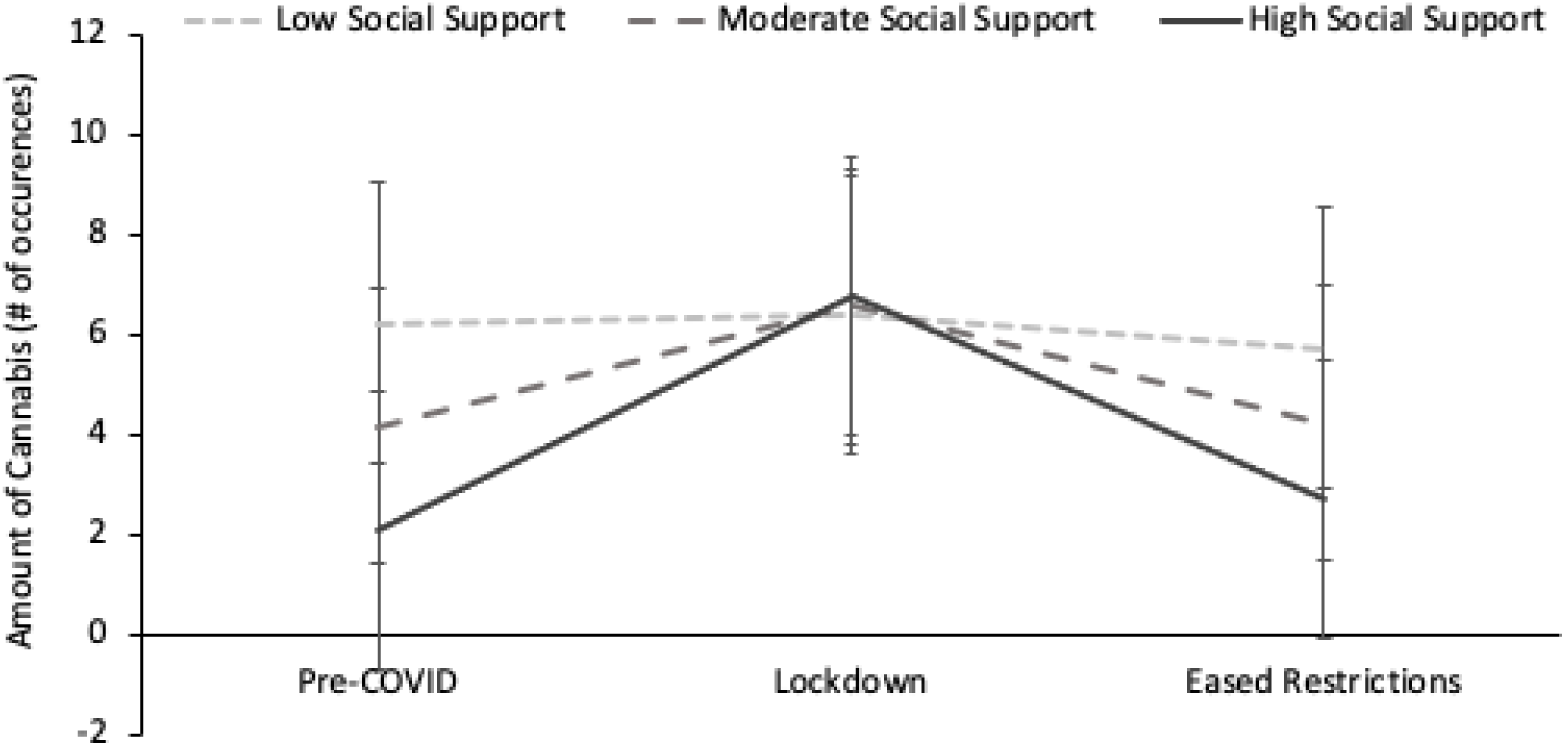
Changing relationship between social support and quantity of cannabis consumption throughout COVID-19 time points. Error bars show standard error of the means.

To follow up these interactions we first examined the relationship between Social Support and quantity of Cannabis use at each time point. Similar to the frequency of Cannabis consumption, Pre-COVID and during Eased Restrictions Social Support negatively predicted the quantity of Cannabis use, all bs > -0.103, 95% CI = [-0.156, -0.050]. However, in contrast to the frequency of Cannabis consumption, during Lockdown Social Support did not predict the quantity of Cannabis use, b = 0.013, 95% CI = [-0.041, 0.067].

Second, we examined differences in the frequency of Cannabis consumption across time points at each level of Social Support. The quantity of Cannabis use remained consistent between Pre-COVID, Lockdown, and Eased Restrictions for individuals with low Social Support, all *t*s < 1.03, all *p*s > .56. Conversely, individuals with moderate or high Social Support increased their quantity of Cannabis use from Pre-COVID to Lockdown, all *t*s > 5.15, all *p*s < .001, and then decreased during Eased Restrictions, all *t*s > 5.10, all *p*s < .001. As with the frequency of Cannabis use this decrease was to the same levels as Pre-COVID, all *t*s < 0.90, all *p*s > .64.

## Discussion

COVID-19 lockdowns provided a unique opportunity to assess the impact of Social Support on alcohol and cannabis use in emerging adults. Compared to Pre-COVID, alcohol use decreased during Lockdown, whereas cannabis use increased. These changes were modulated by perceived social support with Social Support negatively predicting alcohol use and positively predicting cannabis use during Lockdown. These patterns were not maintained during Eased Restrictions. Specifically, overall alcohol use increased, and cannabis use returned to Pre-COVID levels. The influence of social support and changes to social environments suggest that distinct motives underlie alcohol and cannabis use in emerging adults.

The overall decrease in alcohol use during Lockdown in our study likely occurred because emerging adulthood is associated with drinking in social contexts (Schwartz & Petrova, 2019; Grucza, Norberg, & Bierut, 2009). When the pandemic struck, many students moved home, which likely contributed to the decrease in alcohol consumption. Reduced opportunities to engage in social drinking, therefore, may explain the overall decrease in alcohol use in our sample. At the same time, the absence of a university environment, itself, was not associated with decreased substance use as cannabis use increased during Lockdown. Individuals may have substituted alcohol with cannabis during this period, reflecting different motives for using the two substances. More specifically, drinking is socially motivated (e.g., Grant et al., 2007; O’Hara et al., 2015; Van Damme et al., 2013; Vaughan et al., 2009) and there were few opportunities for social interactions during Lockdown. In contrast, cannabis use may be motivated by a desire to reduce stress and anxiety (Bottorff et al., 2009; Hyman & Sinha, 2009; Glodosky & Cuttler, 2020), both of which were elevated during Lockdown. Furthermore, there is evidence that enhancement motives are notably associated with cannabis use in emerging adults (Gette et al., 2021). Taken together, young adults may have increased their cannabis use with the goal of reducing pandemic-related stress and feelings of boredom.

Reductions in alcohol use during Lockdown were highest among individuals with higher social support, suggesting that their alcohol use may be more susceptible to the changes in social contexts that occurred during Lockdown. Moreover, alcohol consumption in young adults is heavily influenced by the drinking behaviors of close friends and social networks (Astudillo et al., 2013; Jamison & Myers, 2008). Consequently, individuals with higher social support may be more prone to these social influences because they presumably have higher quality and quantity of friendships. Although all individuals decreased their quantity of alcohol use during Lockdown, the decrease was larger for those with higher social support, suggesting increased sensitivity to the influence of social environments on drinking. Social support also had less impact on changes in cannabis use among individuals with lower social support: this group used greater amounts of cannabis Pre-COVID but did not increase their use during Lockdown. Baseline stress levels may have been higher in these individuals, making them less sensitive to the additional stress brought on by COVID-19 lockdowns and social isolation. Interestingly, although social support has a protective role against stress and substance use (Li et al., 2021; Ozbay et al., 2007; Rapier et al., 2019; Stone et al., 2012), higher social support did not mitigate the impact of COVID-induced stress on cannabis use. This suggests that the protective role of social support may only be applicable when individuals are not socially isolated. In other words, higher social support individuals may have used cannabis to cope with stress and boredom during social isolation. Taken together, our findings support pre-existing literature that different motives and levels of social support underlie alcohol and cannabis use.

With the emergence of new COVID-19 variants, the world will likely continue to cycle between various extents of social restrictions and lockdowns. Critically, there has been growing concern over the lasting impacts of the pandemic on substance use and mental health (Abbott, 2021; Czeisler et al., 2020; Galea et al., 2020; Singh et al., 2020). Our work adds to these investigations, while showing that changes to alcohol and cannabis use during Lockdown were not sustained when restrictions eased. The rebound in alcohol use during Eased Restrictions provides further evidence that drinking in young adults is socially motivated. Similarly, cannabis use, which increased during Lockdown, returned to Pre-COVID levels during Eased Restrictions, suggesting that individuals with high social support were using cannabis to cope with stress and boredom during social isolation. Regardless of whether changes to substance use during Lockdown were favorable (e.g., decreased alcohol use) or unfavorable (e.g., increased cannabis use), young adults may be resilient to the impacts of COVID-19 lockdowns on substance use.

As with most self-report and retrospective studies, participant calculations of substance use may have been less precise than data collected in real time. The issue was mitigated, at least partially, using the validated TLFB. This measure is widely used to assess retrospective substance use, with several studies showing its validity (Sobell et al., 2001; Martin-Willet et al., 2020; Levy et al., 2004). Measuring cannabis use through ingestions may also have limited precision as the quantity consumed through one ingestion can vary based on the individual and method of consumption. Despite these limitations, we adopted this measure in our study so that our findings would be consistent with previous work using a validated method of quantifying cannabis use (i.e., TLFB). We also acknowledge the difficulty in relating changes in substance use to social restrictions when the timing of these regulations varied across regions in North America. Although there were widespread lockdowns in March 2020, this could have impacted individuals’ substance use throughout the three time points. Moreover, although we recruited participants from both Canada and the United States, the U.S. sample was small (N = 16).

Consequently, we cannot make any definitive conclusions regarding substance use patterns in this group. Nevertheless, the substance use patterns in this group follow the same trends as the rest of the sample. Thus, we decided to include this group as part of the overall sample. The same issues with measuring substance use retrospectively apply to measuring perceived social support retrospectively. However, the MSPSS has good test-retest reliability over several months (Zimet et al., 1988). Additionally, as our sample was primarily composed of university students, findings may not generalize to other emerging adults, or to other age groups. Finally, this sample did not capture any participants who reported identifying as transgender and few (n=5) who identified as gender variant. It is important to acknowledge, therefore, that these findings may not apply to non-cisgender populations.

## Conclusions

Changes to substance use patterns during the COVID-19 lockdowns revealed that different motives may underlie alcohol use compared to cannabis use in emerging adults. Our findings support the idea that socialization in young adulthood plays an important role in alcohol use, especially for individuals with higher social support. Importantly, changes to substance use were not sustained during Eased Restrictions. Several future directions for research are important to clarify the conclusions of this study. Studies using cross-sectional and prospective cohort designs would be most useful to answer the following questions with a higher degree of certainty. First, a direct comparison of alcohol and cannabis use in different living situations should be investigated. Second, the degree that social motivations dictate alcohol use for individuals with higher versus lower social support is important to clarify. Third, more research is needed to investigate whether stress reduction is in fact the primary motivation underlying cannabis use. Finally, studies on substance use throughout the remainder of the pandemic and beyond should be conducted to assess whether young adults are truly resilient to the potential long-term implications of COVID-19 restrictions on substance use.

## Data Availability

Data produced in the present study are available upon reasonable request to the authors

## Acknowledgements

Authors state no acknowledgements.

## Author Contributions

Michelle J. Blumberg: Conceptualization, Methodology, Formal analysis, Investigation, Data Curation, Writing – Original Draft, Writing – Review & Editing, Project administration.

Lindsay A. Lo: Conceptualization, Methodology, Formal analysis, Investigation, Data Curation, Writing – Original Draft, Writing – Review & Editing, Project administration.

Geoffrey W. Harrison: Formal analysis, Data Curation, Writing – Review & Editing, Visualization.

Alison Dodwell: Methodology, Investigation, Writing – Review & Editing. Samantha H. Irwin: Methodology, Investigation, Writing – Review & Editing.

Mary C. Olmstead: Conceptualization, Writing – Review & Editing, Supervision, Funding Acquisition

## Appendix

**Table 1:**
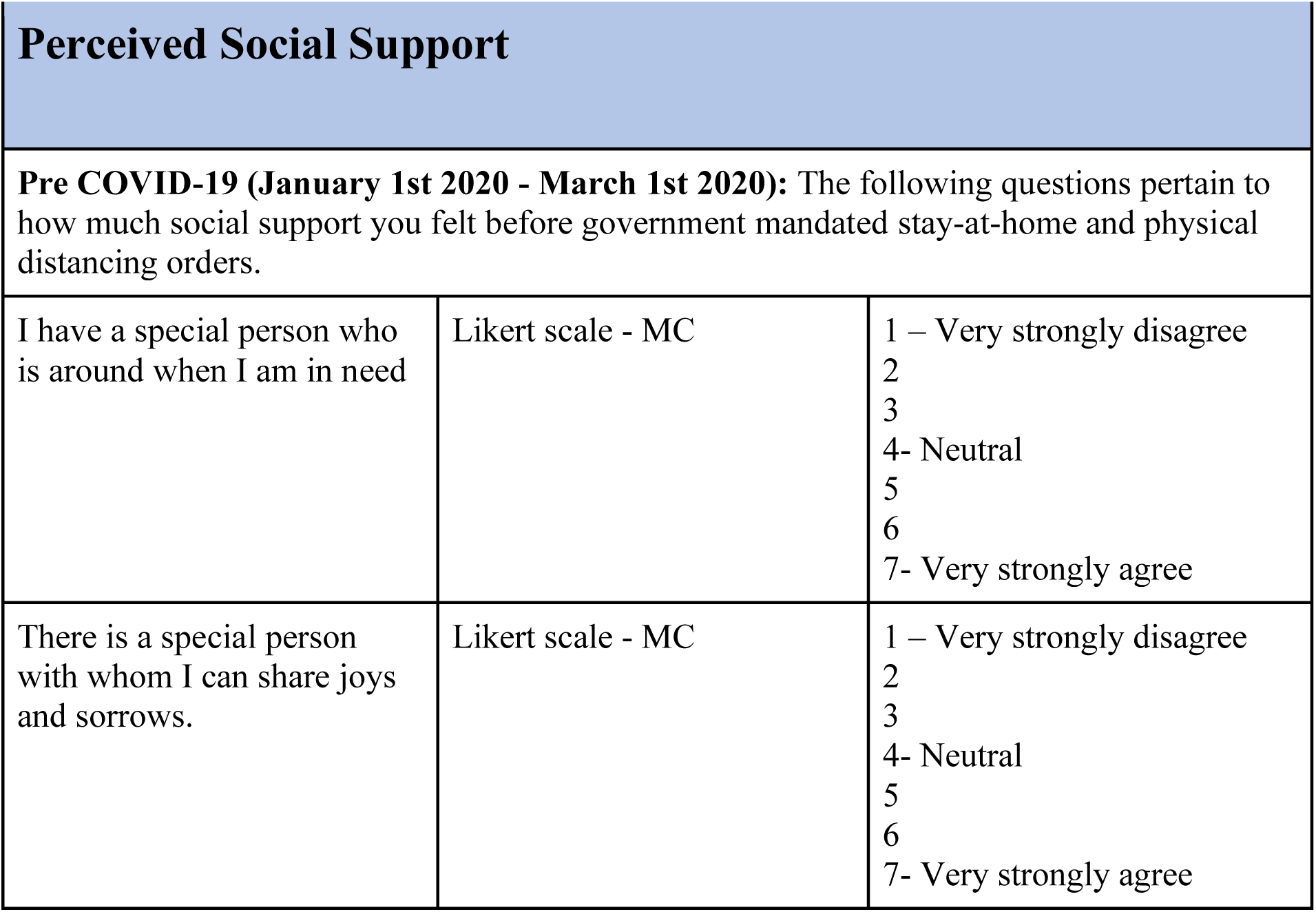
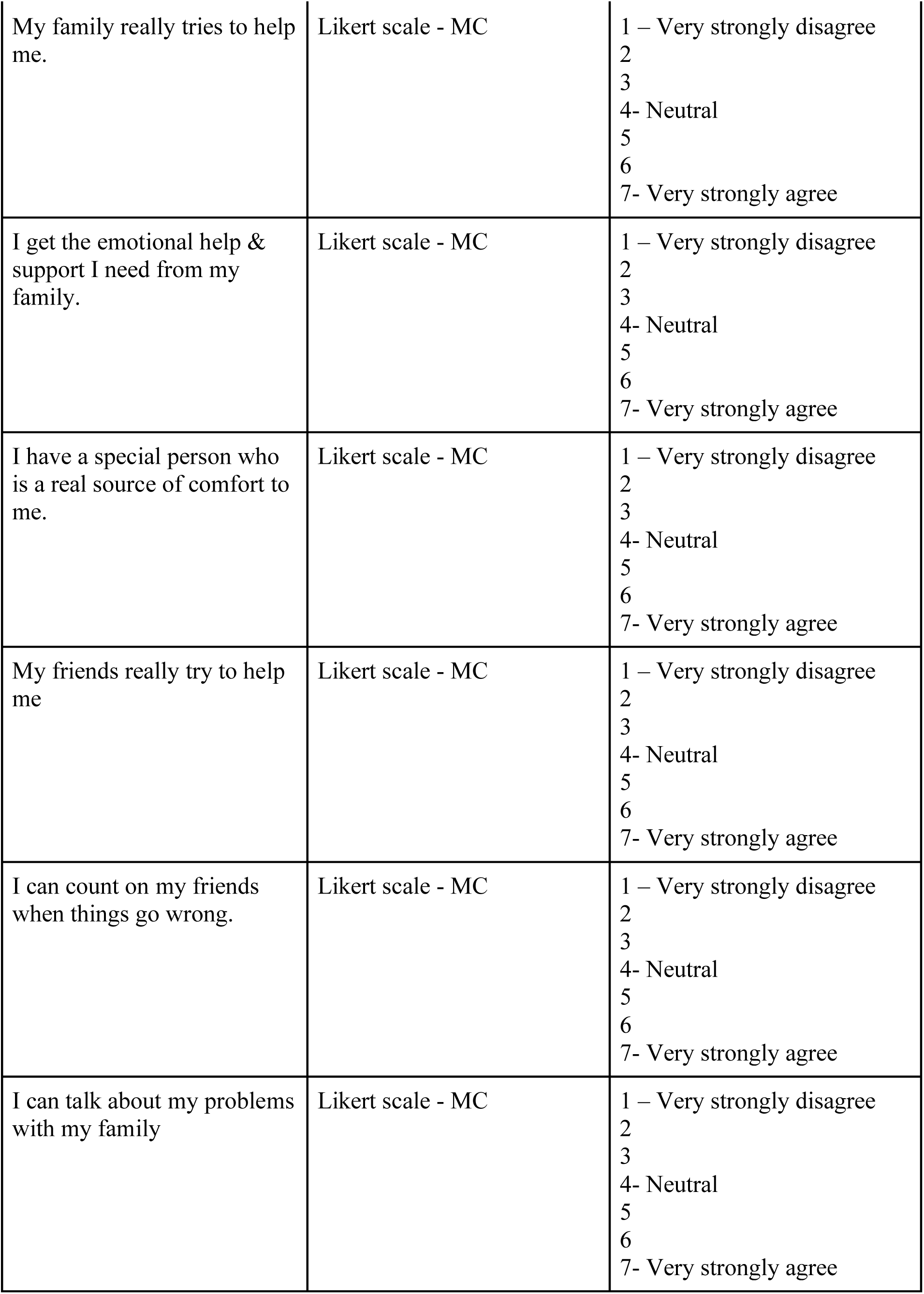
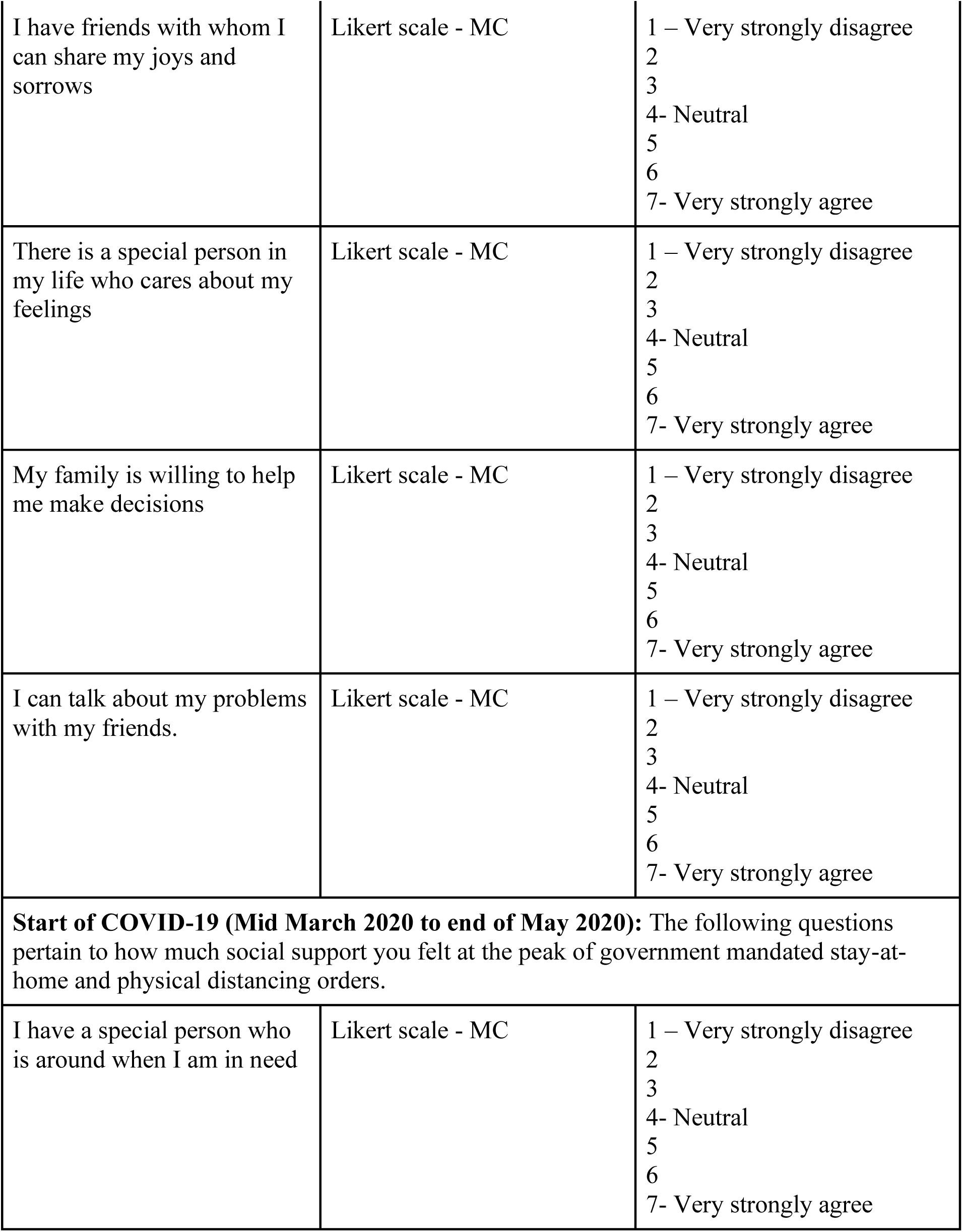
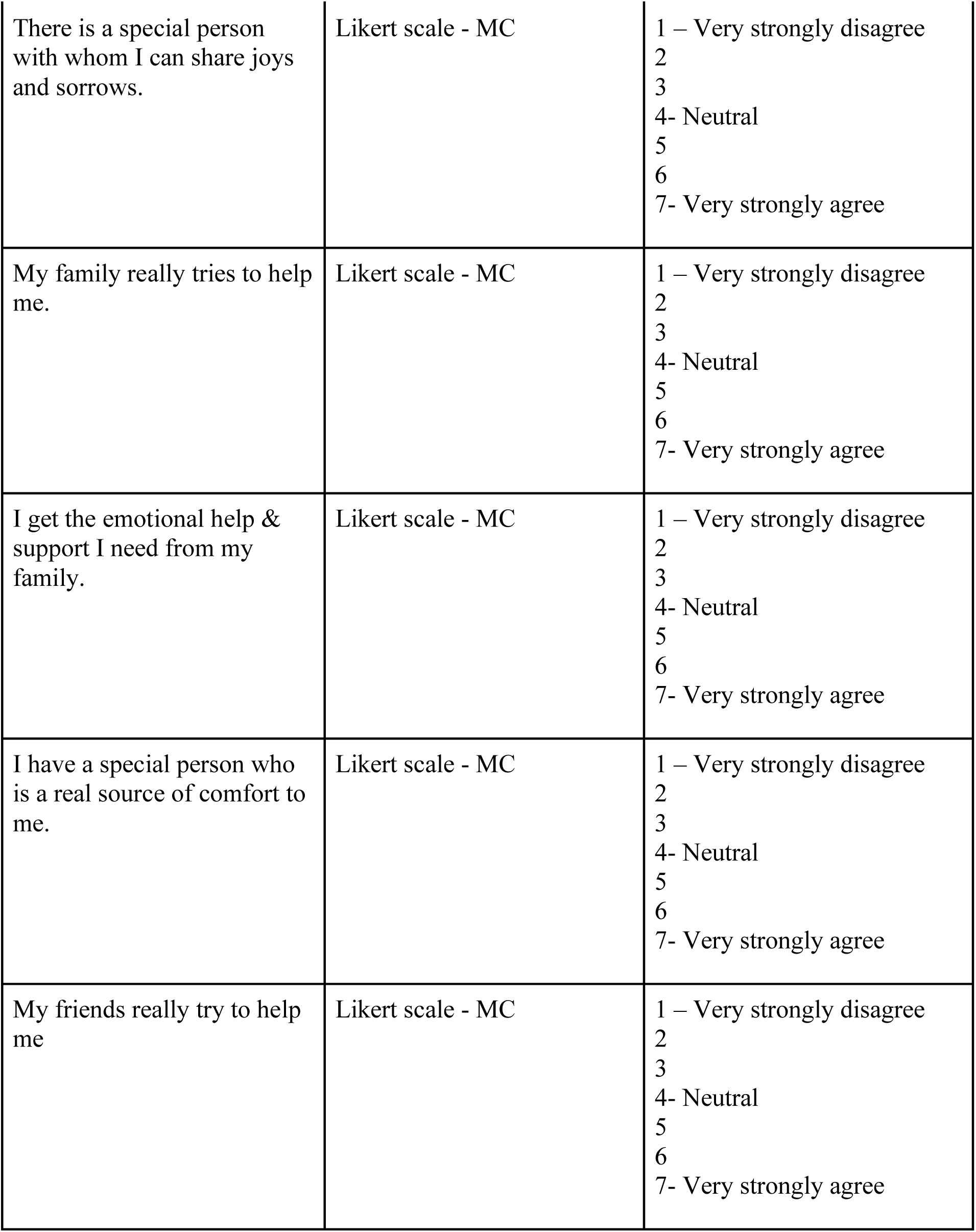
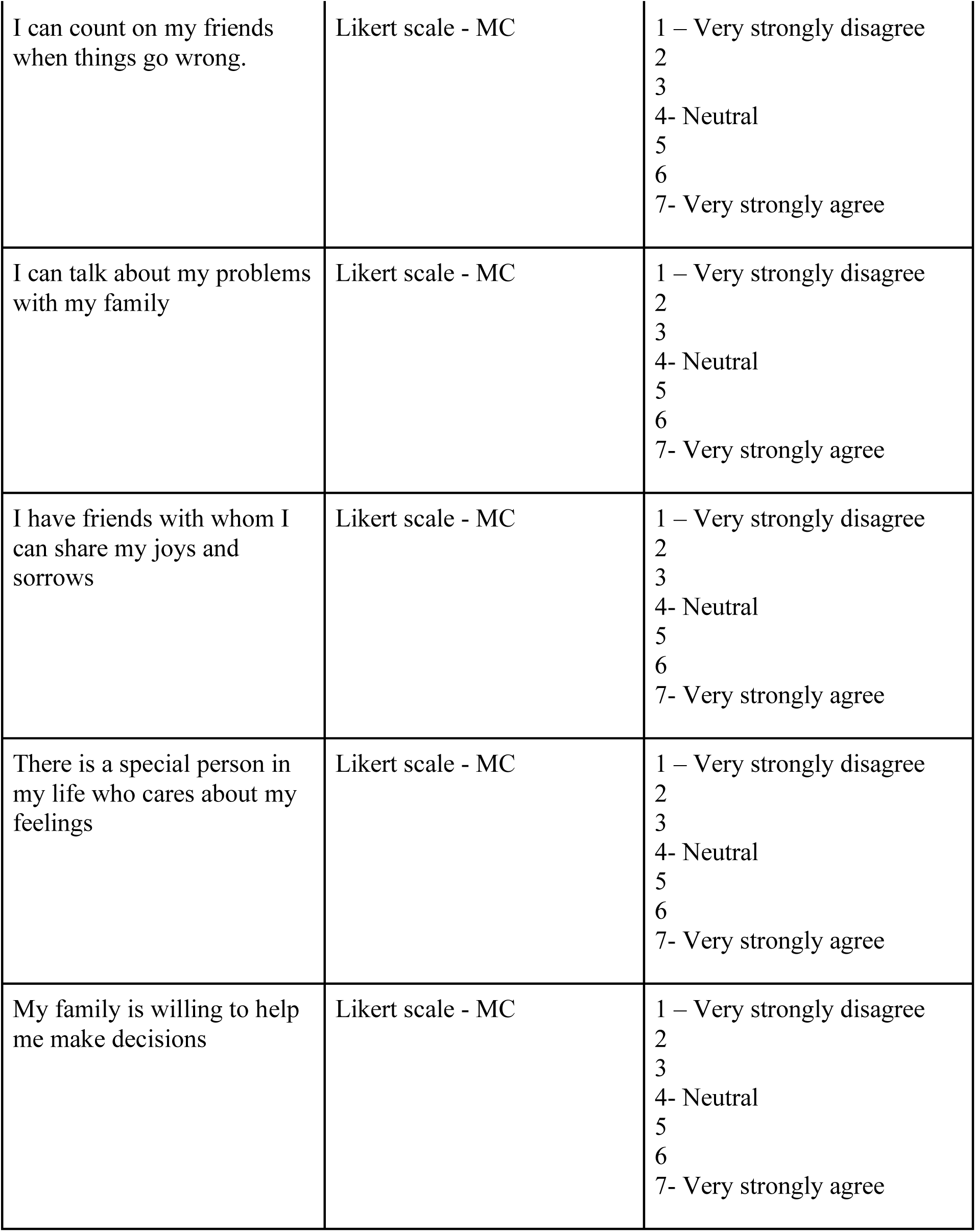
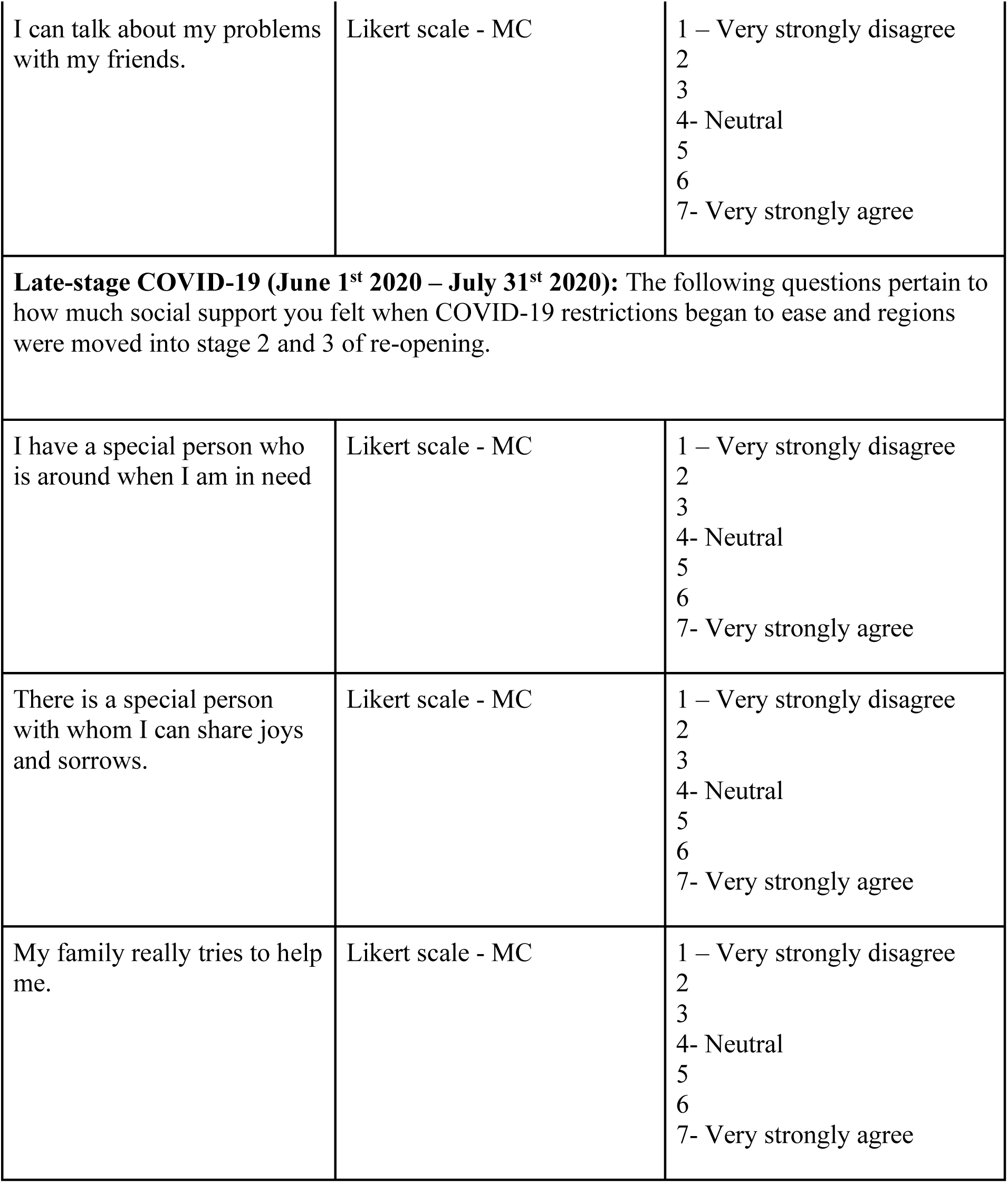
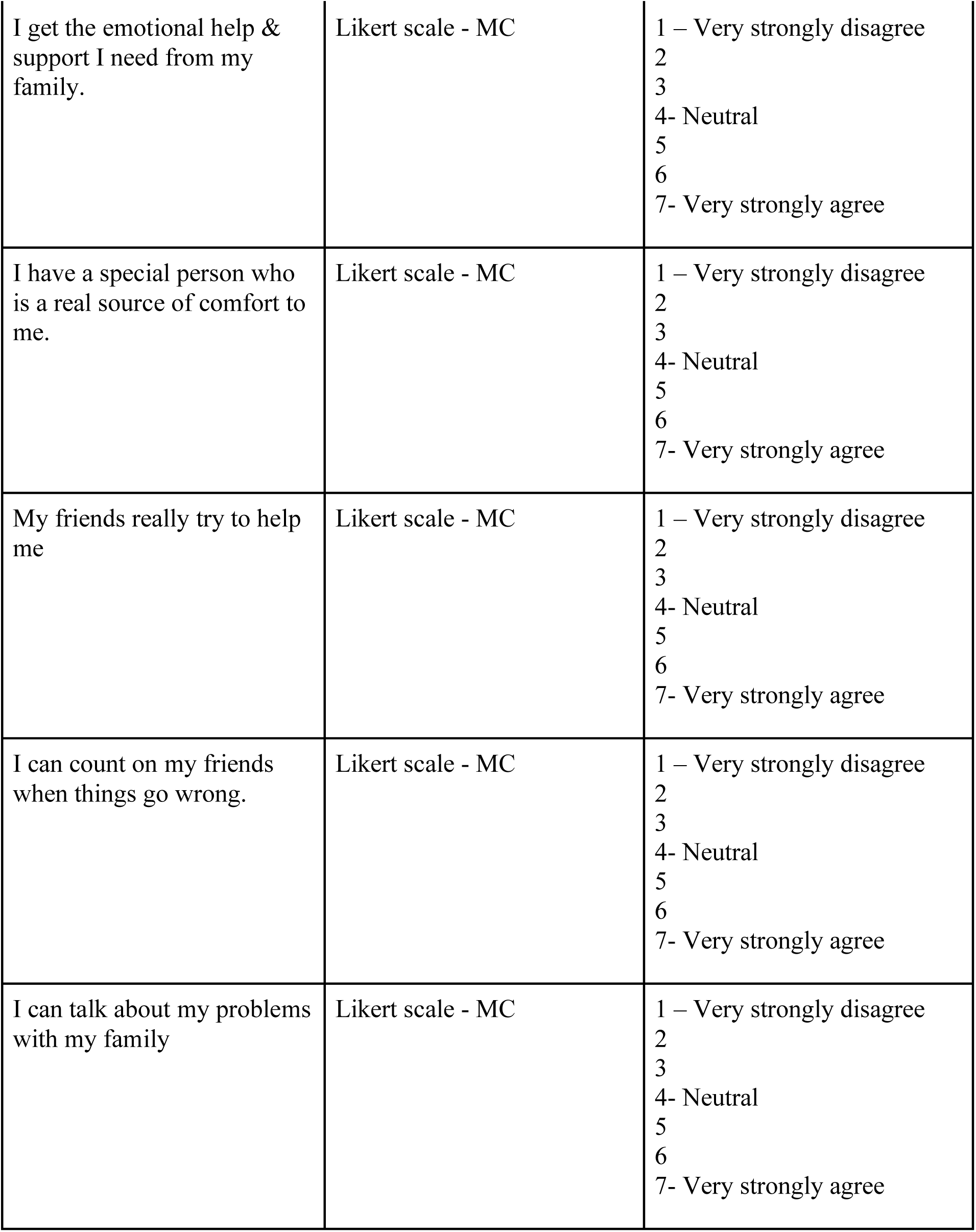
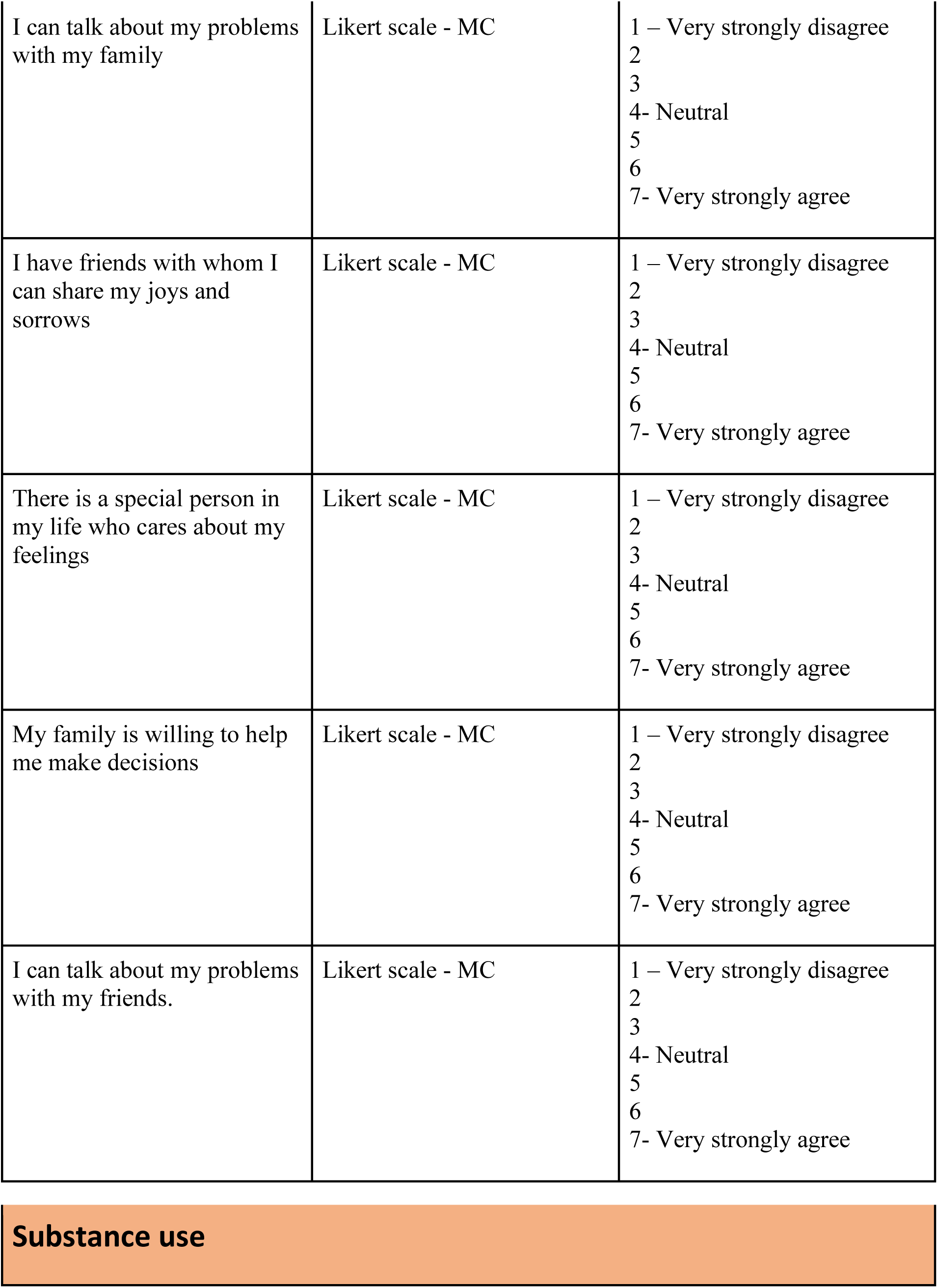
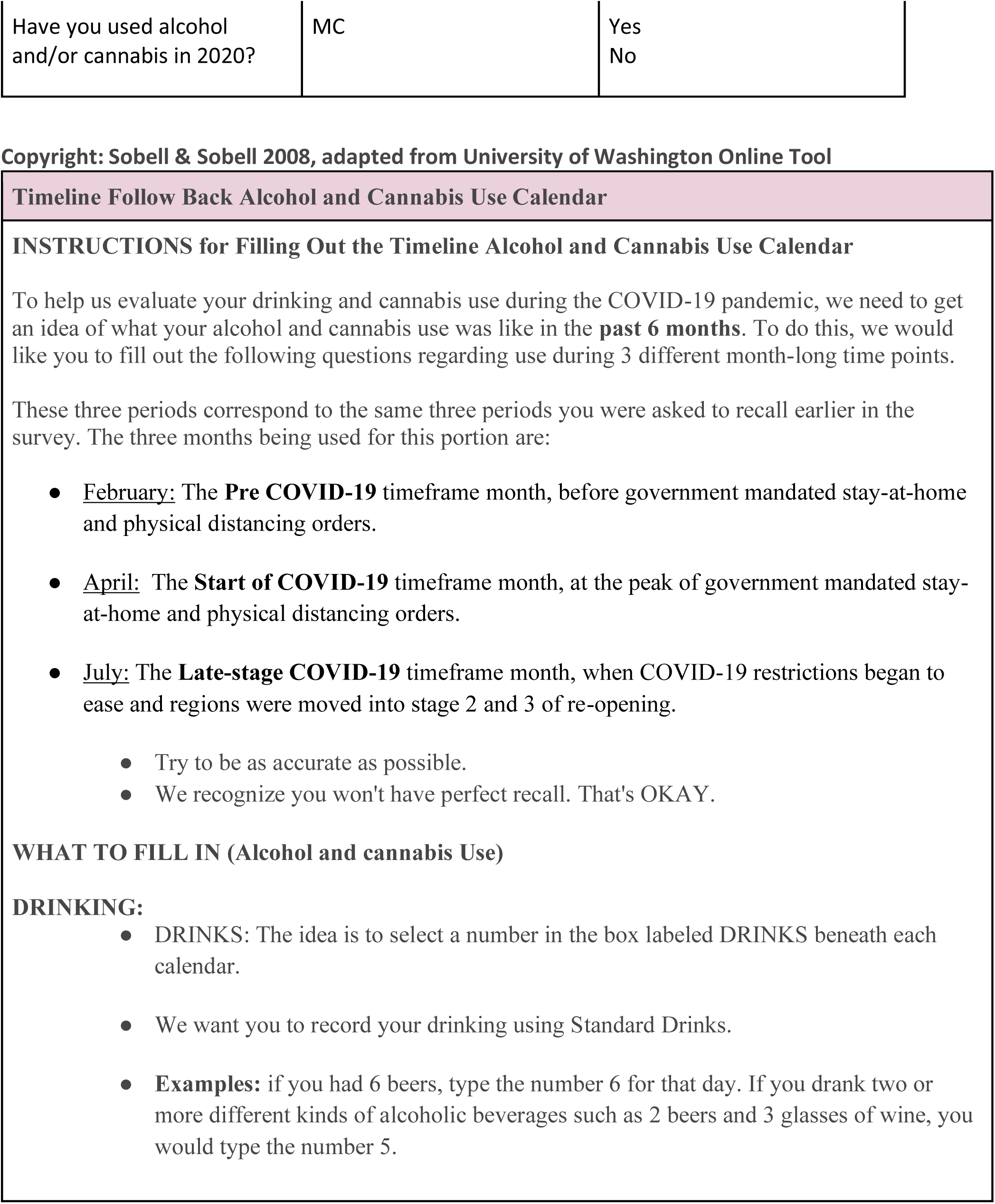
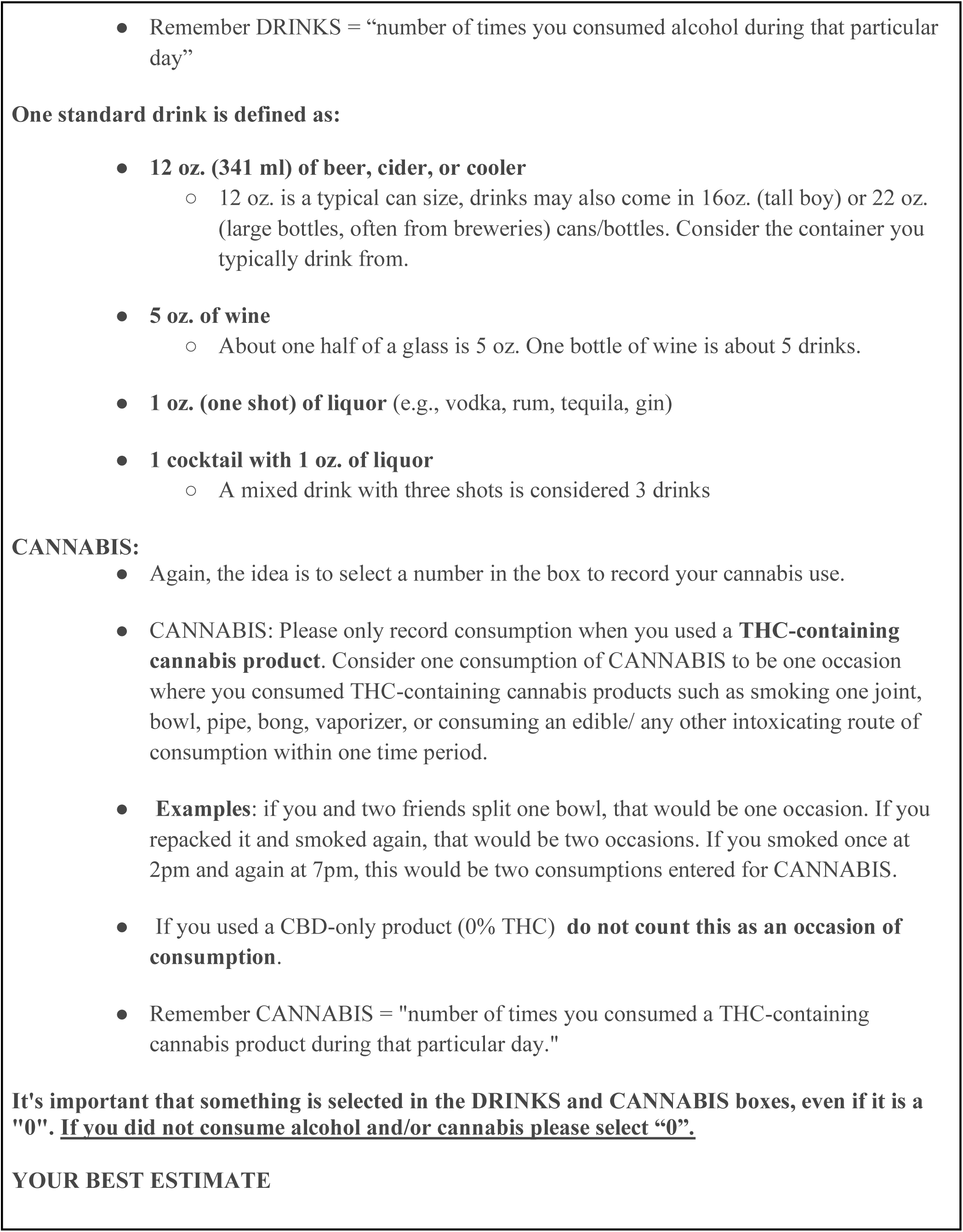
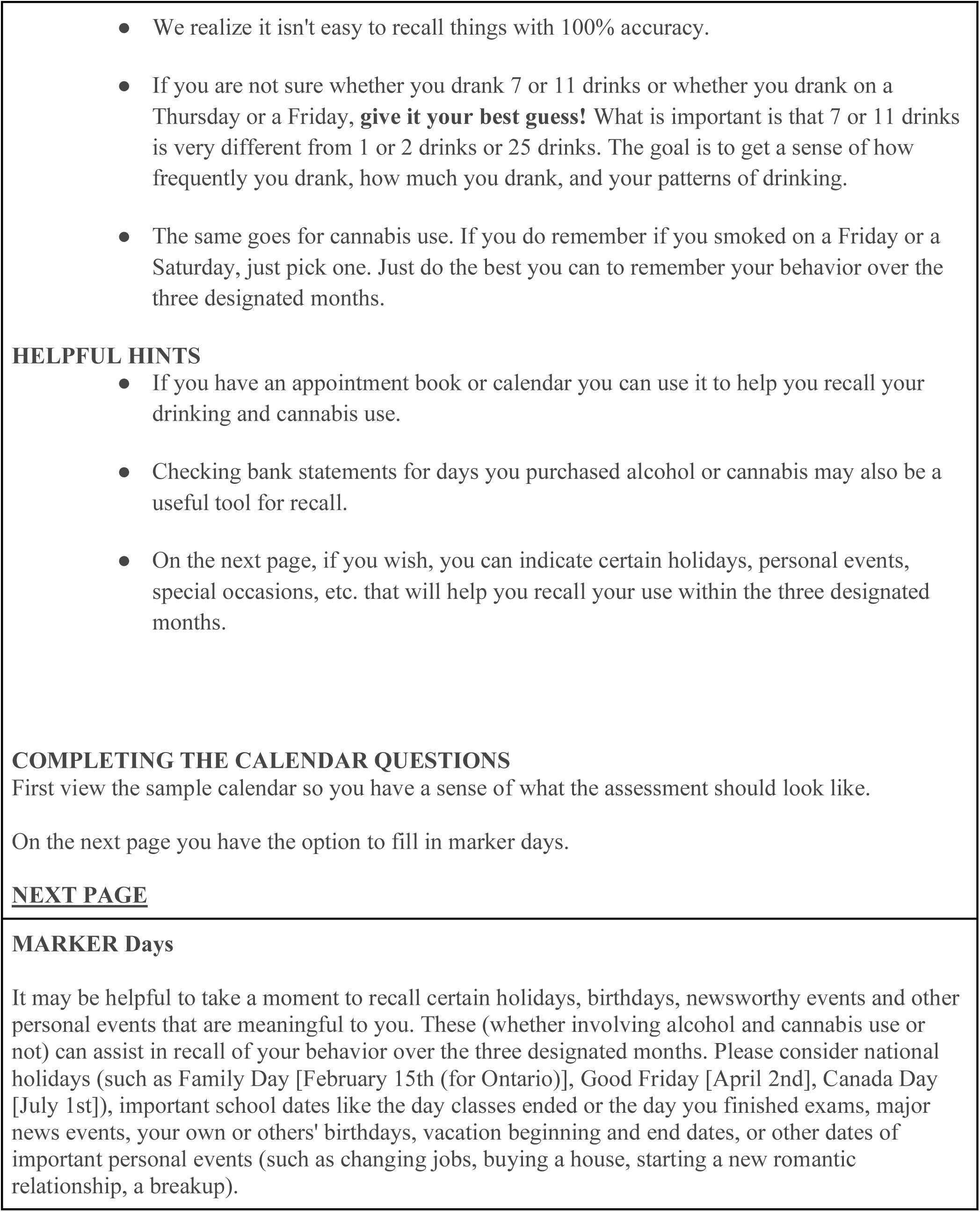
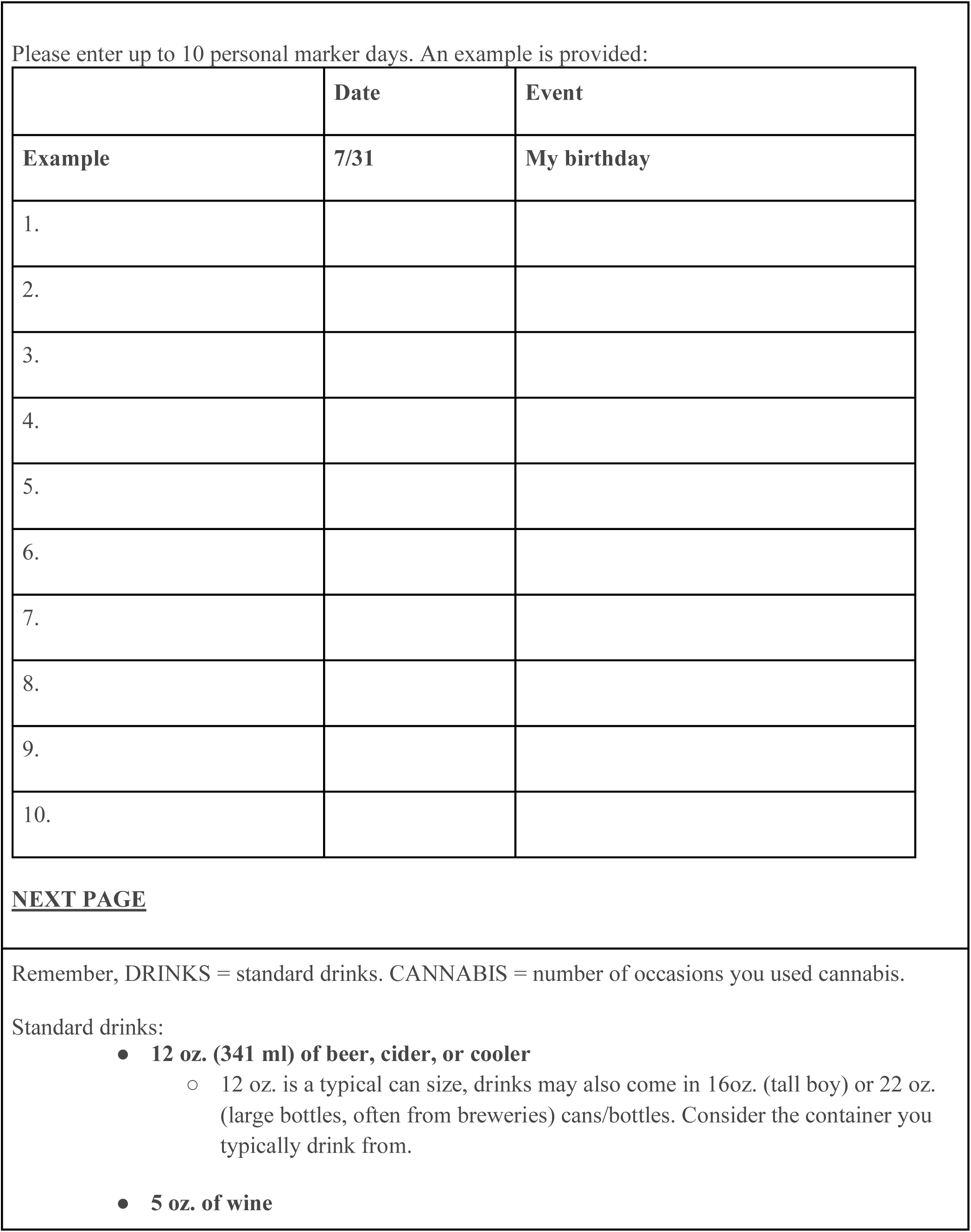
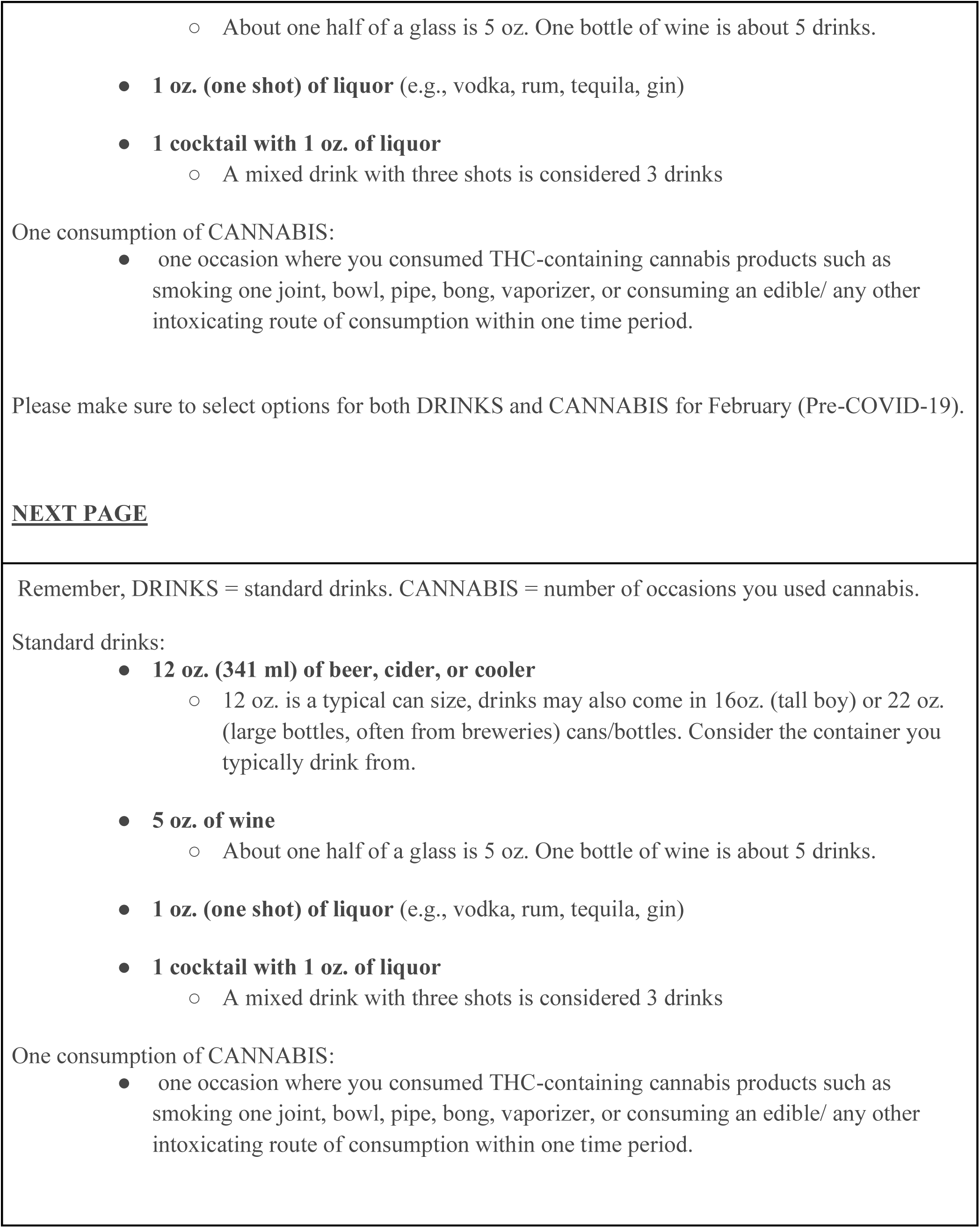
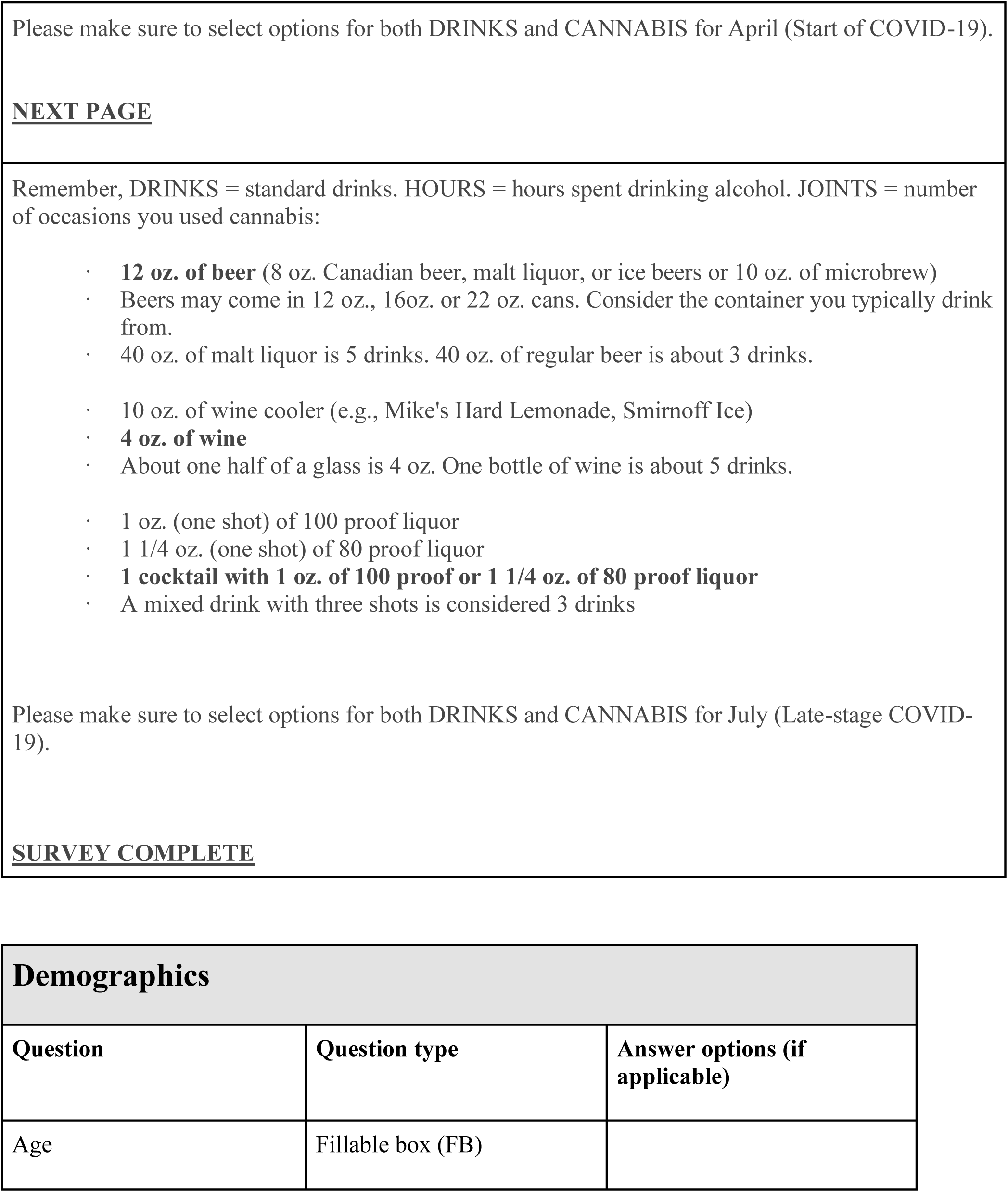
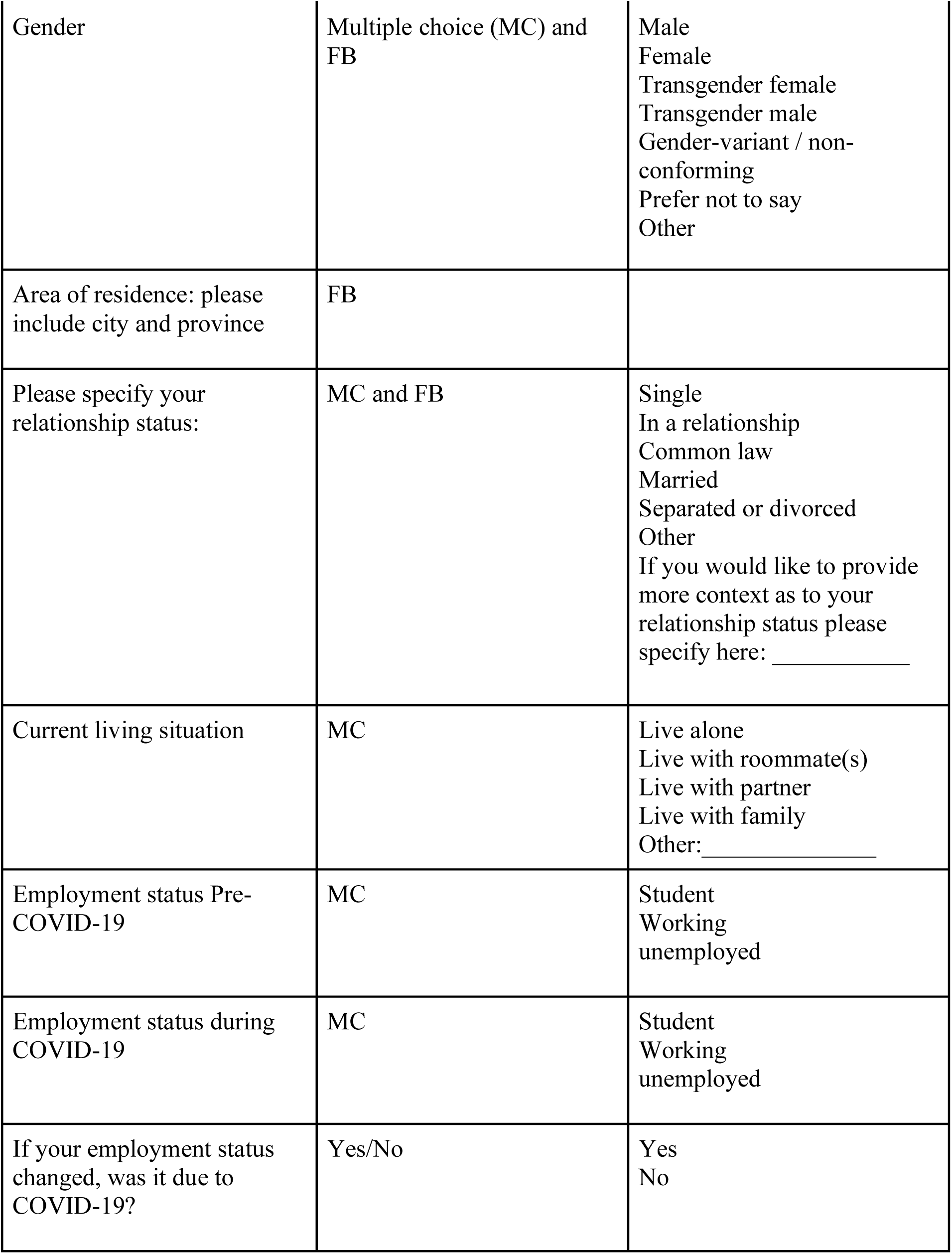
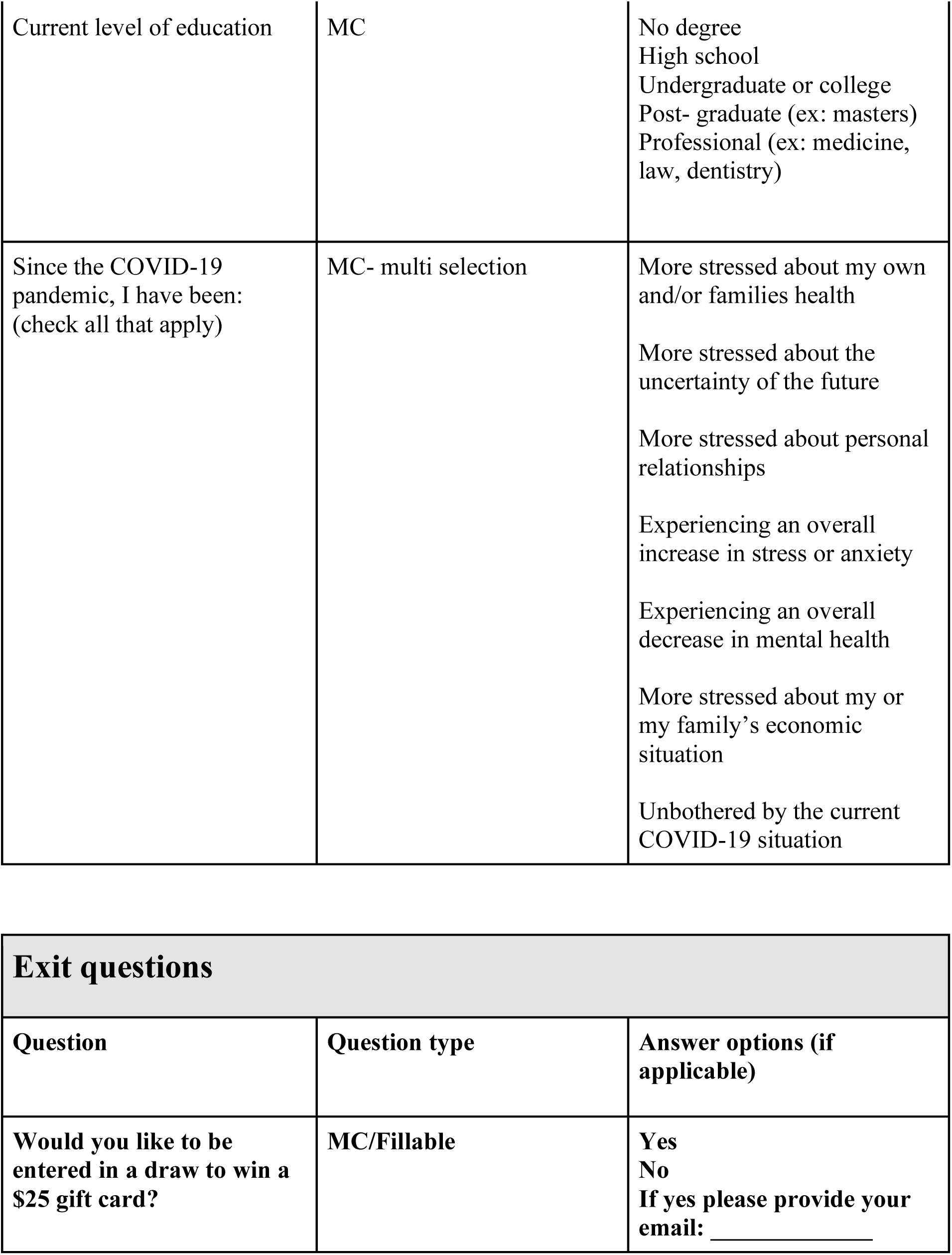
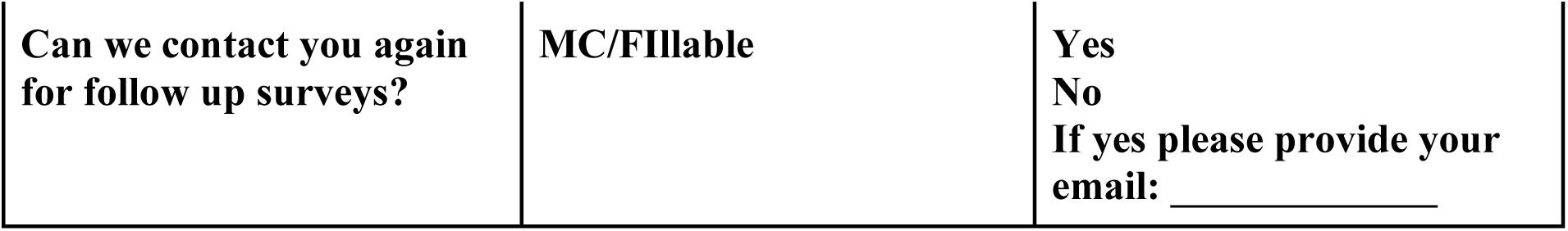
Questionnaire Substance use during the COVID-19 pandemic: A Self-Report Questionnaire The following questions pertain to three time points of the COVID-19 pandemic. The **Pre COVID-19** timeframe refers to the timeframe between January 1st 2020 and March 1st 2020, before government mandated stay-at-home and physical distancing orders. The **Start of COVID-19** timeframe refers to the timeframe between mid-March (approximately March 15th, 2020) and May 31st 2020, at the peak of government mandated stay-at-home and physical distancing orders. The **Late-stage COVID-19** timeframe refers to the timeframe from June 1^st^ 2020 to July 31^st^ 2020, when COVID-19 restrictions began to ease and regions were moved into stage 2 and 3 of re-opening. This includes increasing social circles up to 50 people and the opening of businesses, restaurants, bars, and recreational facilities. Depending on your region these dates may be altered slightly, however, we ask that you answer the following questions with reference to the state of government mandated stay-at-home and physical distancing orders, as listed above in Pre COVID-19, start of COVID-19, and Late-stage COVID-19 timeframes.

